# Application of neighborhood-scale wastewater-based epidemiology in low COVID-19 incidence situations

**DOI:** 10.1101/2022.06.07.22276055

**Authors:** Chamteut Oh, Aijia Zhou, Kate O’Brien, Yusuf Jamal, Hayden Wennerdahl, Arthur R Schmidt, Joanna L. Shisler, Antarpreet Jutla, Arthur R Schmidt, Laura Keefer, William M. Brown, Thanh H. Nguyen

## Abstract

Wastewater-based epidemiology (WBE), an emerging approach for community-wide COVID-19 surveillance, was primarily characterized at large sewersheds such as wastewater treatment plants serving a large population. Although informed public health measures can be better implemented for a small population, WBE for neighborhood-scale sewersheds is less studied and not fully understood. This study applied WBE to seven neighborhood-scale sewersheds (average population of 1,471) from January to November, 2021. Community testing data showed an average of 0.004% incidence rate in these sewersheds (97% of monitoring periods reported two or fewer daily infections). In 92% of sewage samples, SARS-CoV-2 N gene fragments were below the limit of quantification. We statistically determined 10^-2.6^ as the threshold of the SARS-CoV-2 N gene concentration normalized to pepper mild mottle virus (N/PMMOV) to alert high COVID-19 incidence rate in the studied sewershed. This threshold of N/PMMOV identified neighborhood-scale outbreaks (COVID-19 incidence rate higher than 0.2%) with 82% sensitivity and 51% specificity. Importantly, neighborhood-scale WBE can discern local outbreaks that would not otherwise be identified by city-scale WBE. Our findings suggest that neighborhood-scale WBE is an effective community-wide disease surveillance tool when COVID-19 incidence is maintained at a low level.

**Graphical abstract:** 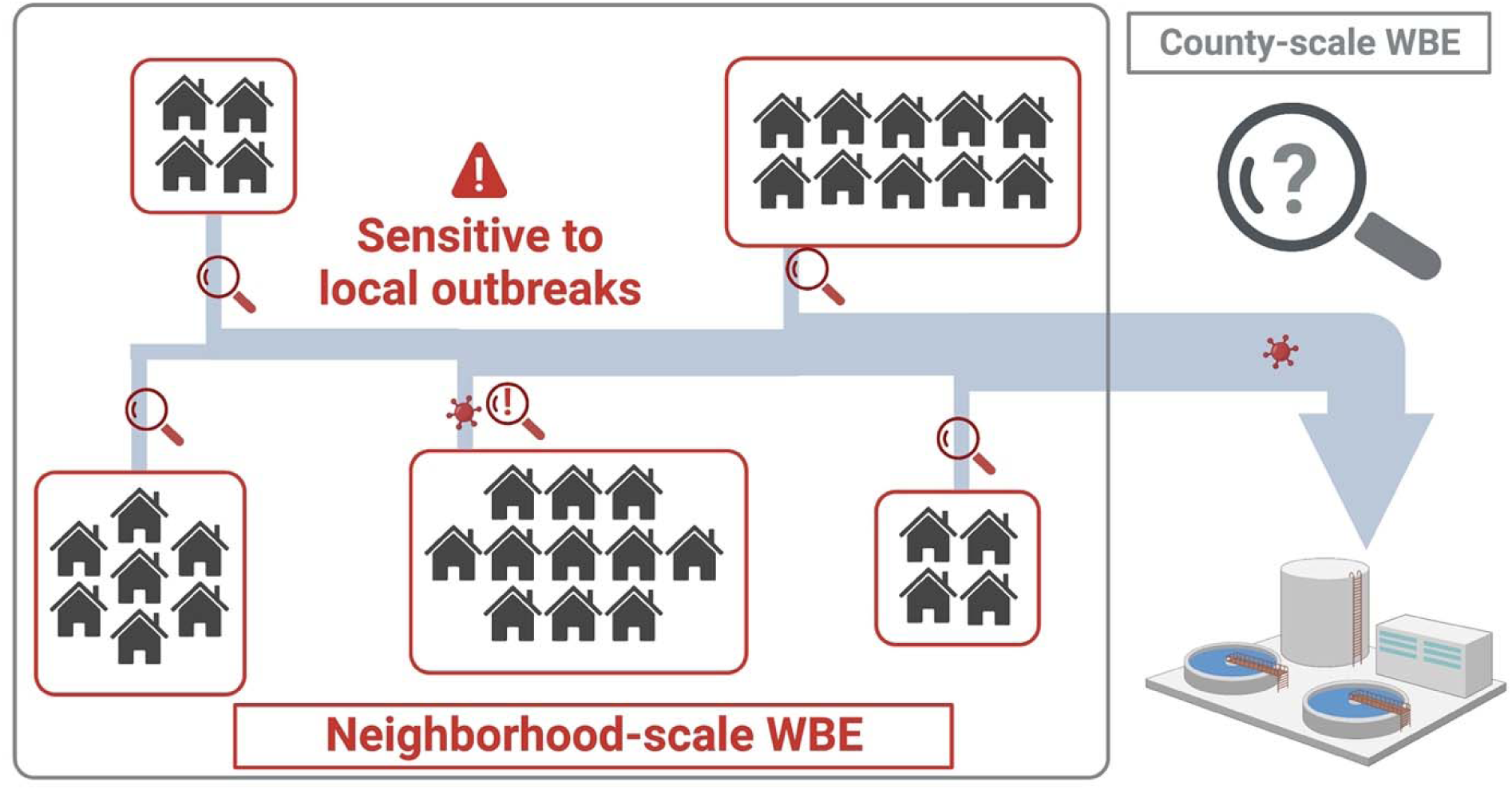

## Introduction

Wastewater-based epidemiology (WBE) has been used as a cost-effective epidemiological tool for community-wide COVID-19 surveillance worldwide (Chen et al., 2020; Hart and Halden, 2020; Sherchan et al., 2021; Tiwari et al., 2021). For example, WBE has been shown to be able to detect SARS-CoV-2 at an 0.01% incidence (1 infection in 10,000 people) when the influence of wastewater treatment plants serving a community of more than 100,000 residents was analyzed (Hata et al., 2021). In addition, the viral genome concentrations in sewersheds may be a better proxy for actual disease incidence than the reported COVID-19 infection cases because reported cases are significantly affected by temporal clinical testing capacity (Xiao et al., 2022). Furthermore, WBE allows for longitudinal wastewater surveillance to predict disease outbreaks in advance of clinical epidemiology because SARS-CoV-2 genes are shed before symptom onset (Wu et al., 2022b).

Large-scale sewersheds (such as wastewater treatment plants serving hundreds of thousands to millions of people) have routinely been surveilled for COVID-19 since early 2020 (Hata et al., 2021; Xiao et al., 2022). However, the large-sized sewersheds may not provide detailed information for region-specific public health measures. For example, social determinants of COVID-19 risk, such as demographics and socioeconomic status, which are regional characteristics (Abrams and Szefler, 2020; Jamal et al., 2022; Upshaw et al., 2021), may not be reflected in the influent wastewater from large sewersheds. The heterogeneity of these social determinants across the sewersheds is a possible explanation for discrepancies between state-wide and county-specific COVID-19 data (Messner and Payson, 2020). WBE also has been applied to small-scale sewersheds, such as buildings on university campuses (Bivins and Bibby, 2021; Gibas et al., 2021; Karthikeyan et al., 2021). At this smaller scale, viruses are detected in wastewater when there are COVID-19 infection cases in the buildings. However, building-scale monitoring is not practical for residential areas with high numbers of single-family homes.

Neighborhood-scale monitoring for COVID-19 has been conducted (Barrios et al., 2021; Spurbeck et al., 2021). While this neighborhood-scale monitoring could allow for a more targeted public health intervention in a specific community, SARS-CoV-2 N gene concentrations presented lower correlation coefficients with COVID-19 infection cases as the catchment populations decreased (Bitter et al., 2022; Rusiñol et al., 2021; Sangsanont et al., 2022). Additionally, the probability of SARS-CoV-2 detection in wastewater samples at a given COVID-19 incidence decreases with a decrease in the population size (Fitzgerald et al., 2021; Wu et al., 2021). Thus, neighborhood scale monitoring strategies are a possible area of improvement.

This study aims to develop a methodology for the neighborhood-scale WBE for community-wide COVID-19 surveillance. We collected sewage samples from seven neighborhood-scale sewersheds (average population of 1,471 people) across Champaign County, IL, USA for eleven months. First, we analyzed the SARS-CoV-2 N gene, S:A570D mutation, and S:P681R mutation. These two mutations are confirmed by in silico analysis and in vitro experiments to be exclusive to the Alpha and Delta variants circulating in IL, USA, during our monitoring period (Oh et al., 2022b). Then, we plotted these data onto the clinical COVID-19 testing data obtained from the neighborhood-scale sewersheds, allowing us to estimate high COVID-19 incidence with WBE data. Our data uncovered local outbreaks that county-scale WBE would not detect and the introduction of SARS-CoV-2 variants of concern to the sewersheds even at low COVID-19 incidence. Our findings suggest that the neighborhood scale WBE will contribute to COVID-19 surveillance, especially when COVID-19 incidence is low.

## Materials and Methods

### Collection of raw sewage composite samples

We selected eleven sampling points of manholes across Champaign County, IL, USA. Among these, four sampling points (C1, C2, C3, and C4) receive sewage discharged from Champaign and Urbana cities, and three sampling points (R1, R2, and R3) receive sewage discharged Rantoul town. The areas of these seven neighborhood-scale sewersheds vary from 0.09 to 1.70 km^2^. The populations in these sewersheds ranged from 853 to 2402 in 2020 (**Fig. 1**). We monitored these seven sewersheds (C1, C2, C3, C4, R1, R2, and R3) from January 2021 to November 2021, during which we collected 254 sewage samples in total. We also collected wastewater from a food processing plant. The amount of human fecal matter released in this wastewater is much smaller than the processing water from slaughtering and meatpacking. For this reason, we used this wastewater for process control, evaluating cross-contamination among the sewage samples during the sample processing.

**Fig. 1.**
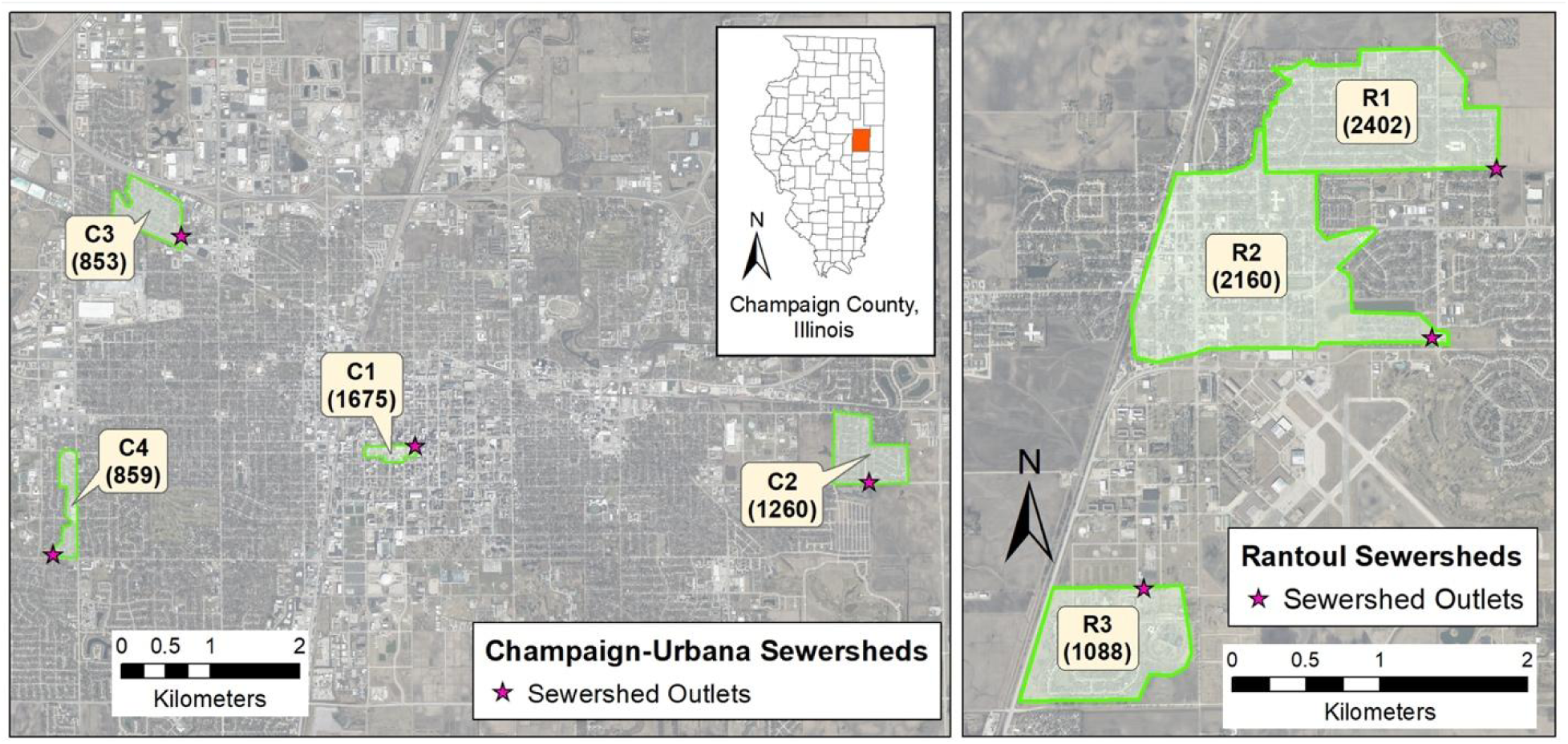
Map of sewersheds in (A) Champaign and Urbana cities and (B) Rantoul town in Illinois. Values in parenthesis are population estimates using block-level US Census 2020 data.

In 2022, we chose another three sampling points in rural areas of Champaign County (S1, S2, and S3), which were less accessible to clinical testing, to supplement community-wide COVID-19 surveillance. Because the sanitary sewer was not connected to most houses in these areas (S1, S2, and S3 served populations of 9094, 3547, and 5547 people, respectively), we collected sewage from local schools that could represent COVID-19 incidence in these areas. It has been shown recently that surveillance at school gave insight into community transmission (Castro-Gutierrez et al., 2022). We also examined wastewater from the Urbana-Champaign Sanitary District (IL, USA), serving 144,097 people living in the cities of Champaign and Urbana and adjacent areas. Although there were two city-scale wastewater treatment plants in this district, we collected wastewater from one of these two plants (City) with the assumption that each wastewater treatment plant represents the entire sewershed. We analyzed 44 samples from the four sewersheds (S1, S2, S3, and City) from January 2022 to March 2022.

We installed autosamplers (Teledyne ISCO, USA) programmed to collect a 1 to 2 L composite sample of sewage pumped at 4-hour intervals for four days. First, the composite samples were transferred to sterile sampling bags (14-955-001, Fisher Scientific, USA), and 20 mL of 2.5 M MgCl_2_ was added to the samples (i.e., final MgCl_2_ concentrations from 25 to 50 mM) to coagulate solids including virus particles (Ahmed et al., 2020b; Oh et al., 2022a). Next, samples were transported on ice to a laboratory at the University of Illinois Urbana-Champaign within three hours. All contact material was sanitized with 10% bleach and 70% ethanol to minimize cross-contamination whenever handling different samples. The minimum recommended meta-information on sewage samples is summarized in **Table S1**.

### Sample processing and viral nucleic acid extraction

Upon arrival at the laboratory, supernatants from each composite sample were discarded. The remaining 35 mL of sewage, in which solid particles were concentrated, were transferred to a 50 mL tube (12-565-271, Fisher Scientific, USA). Next, we added 350 μL of 10^5^ gene copies (gc)/μL of bovine coronavirus (BCoV; 16445-1, Merck Animal Health, USA) to the 35 mL of concentrated sewage. After a 10 minutes incubation at room temperature, samples were centrifuged at 10,000 g for 30 minutes (Sorvall™ RC 6 Plus, Thermo Scientific, USA). Supernatants were then discarded and a portion of the concentrated sludge (100 μL) was transferred to a sterile 1.5 mL tube (1415-2600, USA Scientific, USA). Total RNA and DNA, including SARS-CoV-2 RNA, were extracted from the sludge with QIAamp Viral RNA mini kit (Qiagen, German) following the manufacturer’s procedure with a minor modification. The final volume of extracts was 100 μL. Sewage collection and processing were conducted on the same day, and the RNA samples were stored at −80 L until downstream analysis.

### Analysis of viral genomes

Total RNA was diluted serially 10-fold, and six reverse transcriptase quantitative polymerase chain reaction (RT-qPCR) assays (**Table S2**) were used to analyze samples for the presence of SARS-CoV-2 RNA. We quantified the N gene (i.e., N1 assay with 2019-nCoV RUO kit, Integrated DNA Technologies, USA) to identify a total SARS-CoV-2 RNA concentration. We also analyzed samples for the presence of the S:A570D and S:P681R mutations in the Alpha and Delta variants, as these were dominant variants in our study area during our sample collection (Oh et al., 2022b). For the data quality assurance, we quantified pepper mild mottle virus (PMMOV), Tulane virus (TV), and BCoV, which were used to normalize SARS-CoV-2 to human fecal matter, determine the impact of PCR inhibitors, and calculate SARS-CoV-2 recovery efficiency, respectively.

The details for RT-qPCR assays are summarized in **Table S3,** following MIQE guidelines (Bustin et al., 2009). We analyzed five targets (i.e., N gene, S:A570D, S:P681R, PMMOV, and TV) by Taqman-based RT-qPCR assays. We designed two duplex RT-qPCR assays and one singleplex RT-qPCR to analyze these five targets. We used the first duplex RT-qPCR assay to detect total SARS-CoV-2 RNA (i.e., N gene) and the S:A570D mutation. The second was used to analyze the S:P681R mutation and PMMoV. To validate the two duplex RT-qPCR assays, we compared Cq values of serial dilutions of synthetic RNA/DNA standard controls determined by duplex RT-qPCR and its corresponding singleplex RT-qPCR. We used singleplex RT-qPCR to measure TV RNA. The Taqman-based RT-qPCR started with mixing 5 μL of RNA sample, 5 μL of Taqman Fast Virus 1-step Master Mix (4444432, Applied Biosystems, USA), 1 μL of primers/probe mixture for each assay (i.e., final concentrations of 400 nM for primers and 200 nM for probes), and various volumes of nuclease-free water that fill up the mixture to 20 μL. For example, we added 1 μL of primer/probe for the N gene, 1 μL of primer/probe for the S:A570D mutation, and 8 μL of nuclease-free water to the RT-qPCR cocktail for the first duplex RT-qPCR assay while 1 μL of primer/probe for TV and 9 μL of nuclease-free water to the RT-qPCR cocktail for the singleplex RT-qPCR assay. The PCR cocktail was placed in 384-well plates (4309849, Thermofisher Scientific, USA) and analyzed by QuantStudio 7 Flex (Thermofisher, USA) with a thermal cycle of 5 minutes at 50L, 20 seconds at 95L followed by 45 cycles of 15 seconds at 95L and 60 seconds at 60L. The PCR standard curves were obtained for every RT-qPCR analysis with 10-fold serial dilutions of synthetic RNA or DNA controls, and average PCR efficiencies for RT-qPCR were 92% (**Fig. S1**). Four replicates of nuclease-free water were run for every RT-qPCR analysis to identify the contamination of reagents or cross-contamination among reactions. These non-template controls tested negative in all RT-qPCR analyses represented in our dataset. We also determined the limit of detection (LOD) and limit of quantification (LOQ) for each assay with 20 replicates of serial dilutions of synthetic controls (**Table S4, S5, and S6**), following a previous study (Oh et al., 2022b).

We analyzed BCoV RNA by a SYBR-based RT-qPCR assay. The RT-qPCR mixture for SYBR-based RT-qPCR assay included 3 μL of RNA sample, 0.3 μL of 10 μM forward and reverse primer, 1.275 μL of molecular biology grade water (Corning, NY, USA), 5 μL of 2×iTaq universal SYBR green reaction mix, 0.125 μL of iScript reverse transcriptase from the iTaq™ Universal SYBR® Green One-Step Kit (1725151, Bio-Rad Laboratories, USA). The PCR cocktail was placed in 96-well plates (4306737, Applied Biosystems, USA) and analyzed by an RT-qPCR system (QuantStudio 3, Thermo Fisher Scientific, USA). The RT-qPCR reaction was performed with a thermocycle of 50L for 10 minutes and 95L for 1 minute, followed by 40 cycles of 95L for 10 seconds and 60L for 30 seconds. Melting curves were analyzed while the temperature increased from 60°C to 95°C. The melting curves showed that the primers were specifically bound to the target genome. The SYBR signal was normalized to the ROX reference dye. The cycles of quantification (Cq) were determined by QuantStudio Design & Analysis Software (v1.5.1). The numbers of technical replicates were 4 for synthetic RNA controls and from 3 to 5 for sewage samples. The PCR efficiencies for RT-qPCR were higher than 85% (R^2^>0.99).

Once RNA concentrations in the final extracts were determined, we calculated RNA concentrations in sewage samples using Eqs. 1-4.

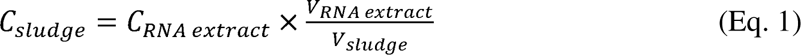

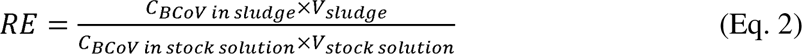

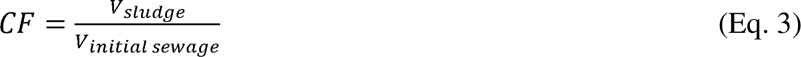

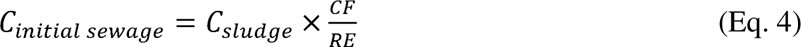

Where C*_RNA extract_*, C*_sludge_*, and C*_initial sewage_*indicate RNA concentration in RNA extract, sludge, and initial sewage, respectively. V*_RNA extract_*, V*_sludge_*, and V*_initial solution_* mean volume of RNA extract, sludge, and initial sewage, respectively. RE and CF represent recovery efficiency and concentration factor, respectively.

### Testing for PCR inhibitors

We evaluated the impacts of potential remaining PCR inhibitors in the RNA extracts by spiking RNA extracts with TV RNA. TV is a calicivirus that infects rhesus monkeys, so the TV should not be present in our collected sewage samples. We added 1 μL of 10^6^ gc/μL TV RNA to 10 μL RNA extracts and 10 μL molecular biology-grade water (Millipore Sigma, Burlington, MA, USA). We assumed that if an RNA extract included PCR inhibitors, a Cq value for TV in the RNA extract would be higher by 1 Cq value or more than in inhibition-free water.

### Presentation of viral genome concentrations

If one of the technical replicates for RT-qPCR is undetermined until 45 PCR cycles, then viral genome concentrations are not quantitatively reliable (Safford et al., 2022; **Fig. S2A**). Hewitt et al. (2022) and **Fig. S2B** showed that positivity/negativity (Eq. 5) is a better indicator for the actual genome concentrations than the Cq values when samples have undetermined RT-qPCR results. We used standard curves to convert Cq values to concentrations when none of the technical RT-qPCR replicates were undetermined. On the other hand, when undetermined data was reported, we used positivity to estimate viral RNA concentrations (Eq. 6).

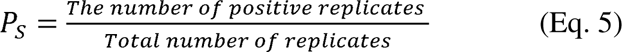

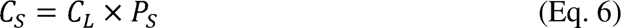

Where C_S_ represents the N gene concentration of samples containing undetermined qPCR results in technical replicates. C_L_ indicates the lowest N gene concentration from samples that do not have undetermined qPCR results. In addition, as suggested in previous studies (Simpson et al., 2021; U.S. CDC, 2022a; Wolfe et al., 2021; Zhan et al., 2022), we presented SARS-CoV-2 RNA concentrations normalized to PMMOV, an indicator for human fecal matter, for comparisons with clinical epidemiology.

### Using BCoV spiked experiments to determine sample composite period

We put 35 mL of sewage sample and 350 μL of 10^5^ gc/μL BCoV particles in a 50 mL sterile tube. The five tubes of the sewage samples containing BCoV were incubated in a water bath where the temperature was maintained at 25L. We collected samples every 24 hours for five days. The number of BCoV RNA was determined by the extraction method and RT-qPCR assays mentioned above. We assumed that the RNA decay follows the first-order decay model (Eq. 7).

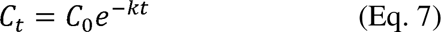

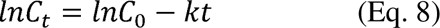

Where C_t_ and C_0_ are the concentration of BCoV RNA at time t and zero, k is the first-order decay rate constant. The rate constant is equal to the slope of ln C and the time graph (Eq. 8), which was determined by the linear least-squares regression model.

### Clinical epidemiology

The Champaign-Urbana Public Health District (CUPHD) provided daily COVID-19 cases with spatiotemporal information for the eight sewersheds (the Urbana-Champaign Sanitary District (UCSD), C1, C2, C3, C4, R1, R2, and R3). Note that there are two wastewater treatment plants in the UCSD, including the one monitored in this study (i.e., City). We assumed that the incidence rates on the City sewershed were the same as that on the entire UCSD sewershed. With the daily COVID-19 cases, we determined the 7-day average and relative infection case. The relative infection case refers to the number of patients actively contributing to sewage viral load. The relative infection case is determined by multiplying temporal daily COVID-19 cases by a virus shedding model (He et al., 2020; Hewitt et al., 2022). We determined the COVID-19 incidence rate for each sewershed by normalizing the COVID-19 occurrence (cases/day) to the corresponding populations.

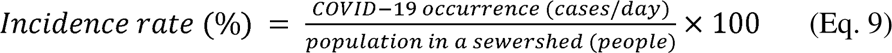

The CDC uses a criterion of 200 people or more COVID-19 cases per 100,000 people for 7 days as one of the metrics to issue a warning of the high level of COVID-19 cases (U.S. CDC, 2022b). According to the virus shedding model, one infected case is assumed to shed viruses 3 days before and 9 days after the symptom onset date. The total virus number of one case shed during this active 13 days equals that of 7.06 people shed on the symptom onset date. Thus, the CDC’s metric is equivalent to a 0.2% incidence rate (Eq. 9).

We analyzed 138 sequences from the Global Initiative on Sharing All Influenza Data (GISAID), which were reported from Champaign County in 2021 using an algorithm, PRIMES developed by Oh et al. (2022b), to determine the temporal prevalence of SARS-CoV-2 variants in Champaign County.

### Statistical analysis

We conducted various statistical analyses to support the credibility of data comparisons. Specifically, BCoV RNA concentrations in sewage with different incubation times were analyzed by Mann-Whitney U Test and a linear least-squares regression model (**Fig. S3**). PMMOV and BCoV RNA concentrations were not normally distributed, so Kruskal Wallis ANOVA was used to compare these data across the seven sewersheds (**Fig. S4** and **Fig. S5**). The Cq values of serial dilutions determined by multiplex and singleplex RT-qPCR assays were compared by F-tests (**Fig. S7**). In addition, N/PMMOV was not normally distributed, so we used Spearman’s rank correlation to compare it to clinical epidemiology (**Table S7**). A receiver operating characteristics (ROC) curve summarizes sensitivity and specificity with a series of N/PMMOV values to indicate COVID-19 incidence rates higher than 0.2%. Sensitivity and specificity were calculated for each N/PMMOV value with Eqs. 10-11.

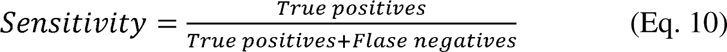

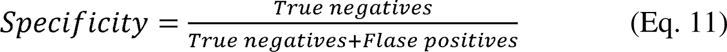

True positives refer to the number of monitoring periods that show high PMMOV and high incidence levels while true negatives refer to the number of monitoring periods showing low PMMOV and incidence levels. False positives indicate the number of monitoring periods that show high PMMOV but low incidence levels while false negatives represent the number of monitoring periods that show low PMMOV but high incidence levels. These statistical analyses were performed using OriginPro (version 2019b). Furthermore, we analyzed the possibility of detecting the SARS-CoV-2 N gene at a given COVID-19 incidence by a binary logistic regression model (**Fig. 5**). This statistical analysis was performed using the *statsmodels* library in Python (version 3.8.8) (Seabold and Josef Perktold, 2010) and the algorithm is deposited at https://github.com/Nguyen205/Logistic_regression.

## Results and Discussion

### Quality assurance and quality control

We first identified an appropriate sample composite period to establish an efficient WBE procedure by spiking samples with known amounts of bovine coronavirus (BCoV) RNA and examining its stability over time. **Fig. S3** used controlled laboratory conditions and showed the concentrations of BCoV RNA as the incubation time increased. The reduction in BCoV RNA concentrations followed first-order decay kinetics (R^2^=1.00) with a first-order decay rate constant of 0.22 day^-1^, which is within the range of previously reported rate constants for SARS-CoV-2 and PMMOV (Roldan-Hernandez et al., 2022). Under these conditions, BCoV RNA concentrations were not significantly different from initial concentrations at four days post-incubation (Mann-Whitney U Test; p>0.05). Ahmed et al. (2020a) found similar results when using SARS-CoV-2. The temperatures measured at our sampling sites were lower than 25L throughout the monitoring period. Therefore, we were assured that coronavirus RNA genomes would remain stable when we collected 4-day composite sewage samples.

PMMOV RNA concentrations were quantified for each composite sample as an indicator that fecal material was present in a composite sample (D’Aoust et al., 2021). The PMMOV RNA concentrations for all collected samples across the seven collection sites are different (Kruskal Wallis ANOVA; p<0.05, **Fig. S4)**. This observation was expected because of the heterogeneity in the sewage composition. Median values of PMMOV RNA concentrations from the seven sewersheds ranged from 10^6.41^ (C2) to 10^7.05^ gc/L (R1), which were similar to those previously reported (Feng et al., 2021). Since PMMOV was detected from all collected sewage samples, our sampling method successfully contained human fecal matter from sewer distribution systems.

We used the recovery efficiency for BCoV to evaluate the sample processing procedures (e.g., sludge concentration and RNA extraction). The mean, median, minimum, and maximum recovery efficiencies across the seven locations were 3.4%, 1.5%, 0.02%, and 64%, respectively. The recovery efficiencies differed significantly (Kruskal Wallis ANOVA; p<0.05, **Fig. S5**). A wide range of recovery efficiency was also reported previously (Feng et al., 2021), and the differences were probably attributed to different sewage characteristics. Pecson et al. (2021) investigated the reproducibility and sensitivity of 36 virus concentration methods performed by 32 laboratories across the U.S. They suggested a threshold of 0.01% of recovery efficiency for the exclusion of poorly processed samples. Based on this criterion, none of the samples were screened out due to the low quality of sample processing.

Sewage samples routinely contain PCR inhibitors (Rački et al., 2014). Thus, it is essential to determine the impact of remaining PCR inhibitors in the final RNA extracts. We conducted experiments to determine the presence of RNA inhibitors by spiking samples with Tulane Virus (TV) RNA. Results are shown in **Fig. S6**. We found that all ΔCq values were within 1.0, confirming that PCR inhibitors’ impacts in our RNA samples were negligible after 10-fold dilution.

We used multiplex RT-qPCR assays to quantify the SARS-CoV-2 N gene, the S:A570D mutation for the Alpha variant, the S:P681R mutation for the Delta variant, and PMMOV replication gene in the final RNA extracts to save samples, reagents, and time. **Fig. S7** shows that the Cq values from singleplex and multiplex RT-qPCR were not significantly different for the two sets of multiplex RT-qPCR assays (F-test; p>0.05). Thus, the fluorescence from FAM dye did not affect the HEX channel and vice versa. For the final negative control, we applied the same procedures, including sample collection, processing, and analysis, to wastewater discharged from a food processing plant and sewage samples. We did not detect a SARS-CoV-2 RNA signal (i.e., Cq values were undetermined by 45 cycles of RT-qPCR) from any of the food processing wastewater samples throughout the monitoring period. This finding suggests no cross-contamination among samples from different sites occurred, so it can be assumed that the positive Cq values were true positives.

### Comparisons of clinical epidemiology and wastewater-based epidemiology data

WBE has been evaluated based on the goodness of fit to clinical epidemiology determined by statistical models such as a Pearson correlation coefficient or a Spearman rank correlation coefficient (Barua et al., 2022; Li et al., 2021), depending on the data normality. Similarly, we compared N/PMMOV ratios to COVID-19 incidence rates determined by three epidemiological parameters: daily COVID-19 cases (Xiao et al., 2022), 7-day average COVID-19 cases (Catherine Hoar et al., 2022), and relative infection COVID-19 cases (Hewitt et al., 2022). Then, we evaluated the goodness of fit by Spearman rank correlation (**Table S7**) between the N/PMMOV ratios and these epidemiological parameters. We found that the relative infection cases yielded higher correlation coefficients than daily cases and 7-day averages at six sewersheds (C1, C2, C3, C4, R1, and R2). Therefore, we used relative infection cases to determine the COVID-19 incidence rate.

The correlation coefficient between COVID-19 incidence rate and N/PMMOV ranged from 0.08 (R3) to 0.72 (C1) across the seven sewersheds (C1, C2, C3, C4, R1, R2, and R3) (**Table S7**). Overall, the correlation coefficients showed a considerable variation among the sewersheds. There are several possible explanations for the lack of uniform correlations.

First, the average COVID-19 incidence rate was 0.004% throughout the monitoring period, equivalent to only a few daily COVID-19 cases in neighborhood-scale sewersheds serving populations of around 1,400 people. **Fig. 2A** shows that no COVID-19 patients were reported at the seven sewersheds for 221 days out of 301 days of average monitoring periods (i.e., 73.4%). One, two, and three or more patients were reported at 18.7, 4.9, and 3.0%, respectively. Individual patients have significant differences in clinical symptoms, viral shedding, and access to COVID-19 tests. For example, Ke et al. (2022) found substantial person-to-person variations in viral load and virus shedding duration among COVID-19 patients. A time lag between symptom onset and testing date also varies because people in different locations have different access to COVID-19 testing. The fact that the different time lags significantly impact the correlation to the WBE data is demonstrated by the highest correlation coefficient (0.72) at location C1. Most residents of location C1 were undergraduate students enrolled at the University of Illinois Urbana-Champaign, which required students to undergo routine COVID-19 testing. For example, graduate students were required to get tested once a week until August 2021. Undergraduate students were required to get tested twice a week until May 2021 and once a week from May to August 2021. While the mandatory COVID-19 testing was lifted for fully vaccinated individuals in September 2021, many residents in campus town still got weekly testing. Therefore, compared to other sites, the reported COVID-19 cases at C1 were much closer to the actual infection cases. The individual heterogeneity in clinical symptoms, including virus shedding and access to COVID-19 tests, will become more significant as the number of COVID-19 cases decreases.

**Fig 2.**
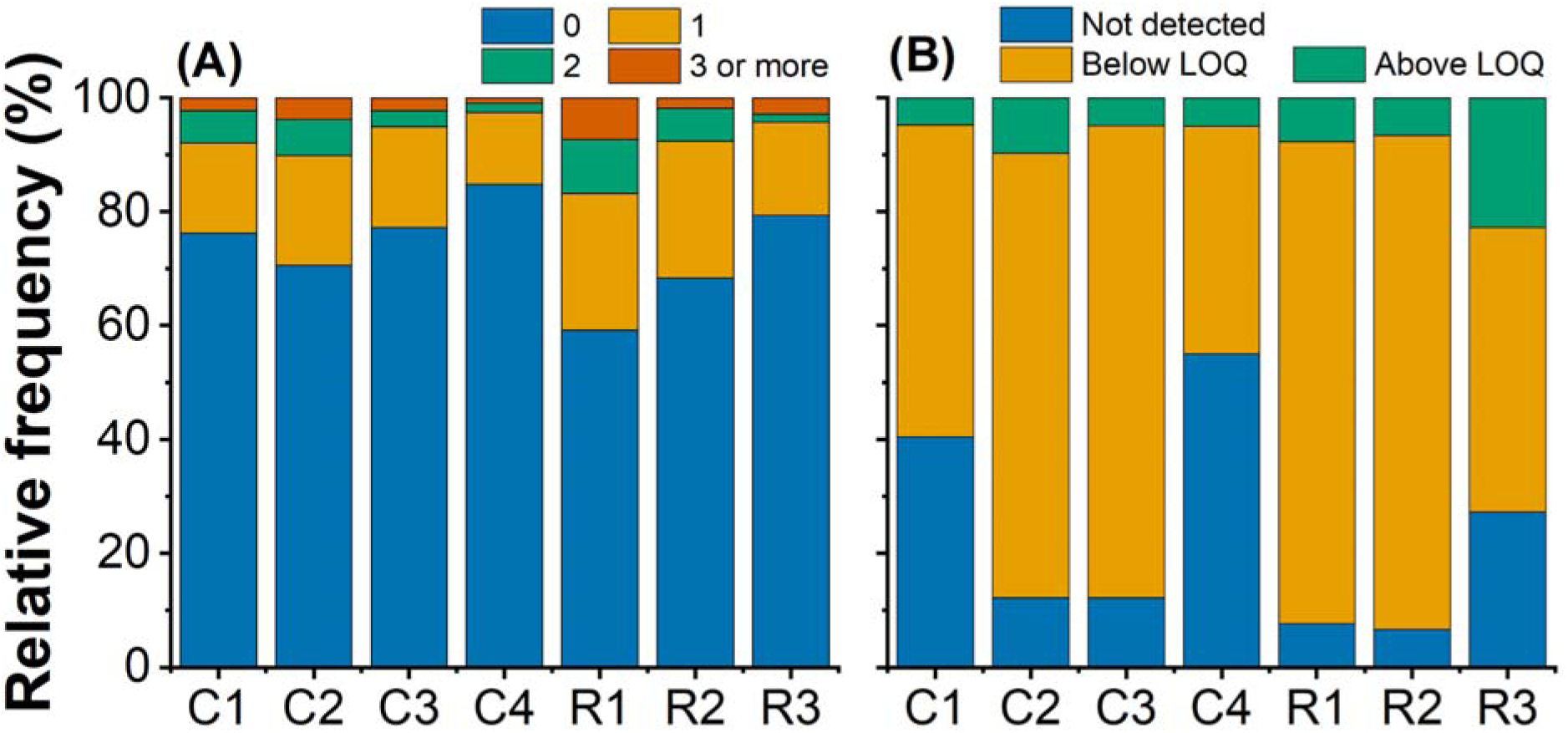
Stacked histograms for (A) COVID-19 daily cases in sewersheds and (B) SARS-CoV-2 concentrations in sewage.

Second, SARS-CoV-2 RNA was present at a low concentration in sewage samples. The SARS-CoV-2 N gene concentrations are summarized in three groups (**Fig. 2B)**; the N gene was not detected; the N gene was detected in at least one of the qPCR replicates, but the concentrations were below LOQ; and the N gene concentrations were above the LOQ. We found that the N gene was detected in 77% of the sewage samples, which agrees with the detection rate of composite wastewater samples determined by meta-analyses (a mean of 70% and a 95%-CI of [47%; 94%]) (Mantilla-Calderon et al., 2022). However, 92% of sewage samples showed concentrations below the LOQ. By definition, the concentrations below LOQ are not quantitatively reliable and may differ from the actual virus concentrations. Therefore, we concluded that the unclear relationship between the WBE and the clinical epidemiological data was attributed to a low number of COVID-19 cases in sewersheds and low SARS-CoV-2 concentrations in sewage.

### Application of wastewater-based epidemiology to neighborhood-scale sewersheds with low COVID-19 incidence rates

We categorized WBE (i.e., N/PMMOV) and clinical epidemiological data (i.e., incidence rate) into two groups and compared the two epidemiological data sets. Firstly, 200 or more new COVID-19 cases per 100,000 populations in the past 7 days is one of the metrics that the Center for Disease Control and Prevention (CDC) uses to decide on the high-risk COVID-19 transmission (U.S. CDC, 2022b), which is equivalent to a 0.2% incidence rate (Eq. 9). We adopted the threshold of 0.2% to categorize incidence rates into high and low levels. Monitoring periods with incidence rates higher or lower than the threshold of 0.2% are shaded by red and blue in **Fig. 3**. The incidence rates were higher than the threshold 158 times (i.e., high incidence rate) and lower 95 times (i.e., low incidence rate) in the seven sewersheds. We investigated if WBE data can estimate these high and low COVID-19 incidence periods. Thus, secondly, we statistically determined a threshold of N/PMMOV, grouping them into a high or low level. The ROC curve summarizes sensitivity and specificity with a series of thresholds of N/PMMOV (**Fig. 4**). We chose 10^-2.6^ as the unbiased optimal threshold of N/PMMOV because it gives the highest geometric mean (G-mean) (Gour and Jain, 2022). The threshold of N/PMMOV is the horizontal dashed line in **Fig. 3**. The N/PMMOV values were higher than the threshold 174 times and lower 79 times. With the threshold of 10^-2.6^, we can predict the COVID-19 incidence rate higher than 0.2% with a sensitivity of 0.82 and a specificity of 0.51. Note that there were probably unreported COVID-19 cases in our sewersheds. For example, the CDC estimates that actual COVID-19 infections are 6 to 24 times higher than reported cases (Havers et al., 2020). These unreported COVID cases might have accounted for the false-positive scenarios where the high N/PMMOV was detected from sewage samples while the COVID-19 incidence rate was low. If so, the true specificity becomes higher than 0.51. These findings suggest that the threshold of N/PMMOV of 10^-2.6^ can be used to indicate the high risk of COVID-19 transmission in the neighborhood-scale sewersheds.

**Fig. 3.**
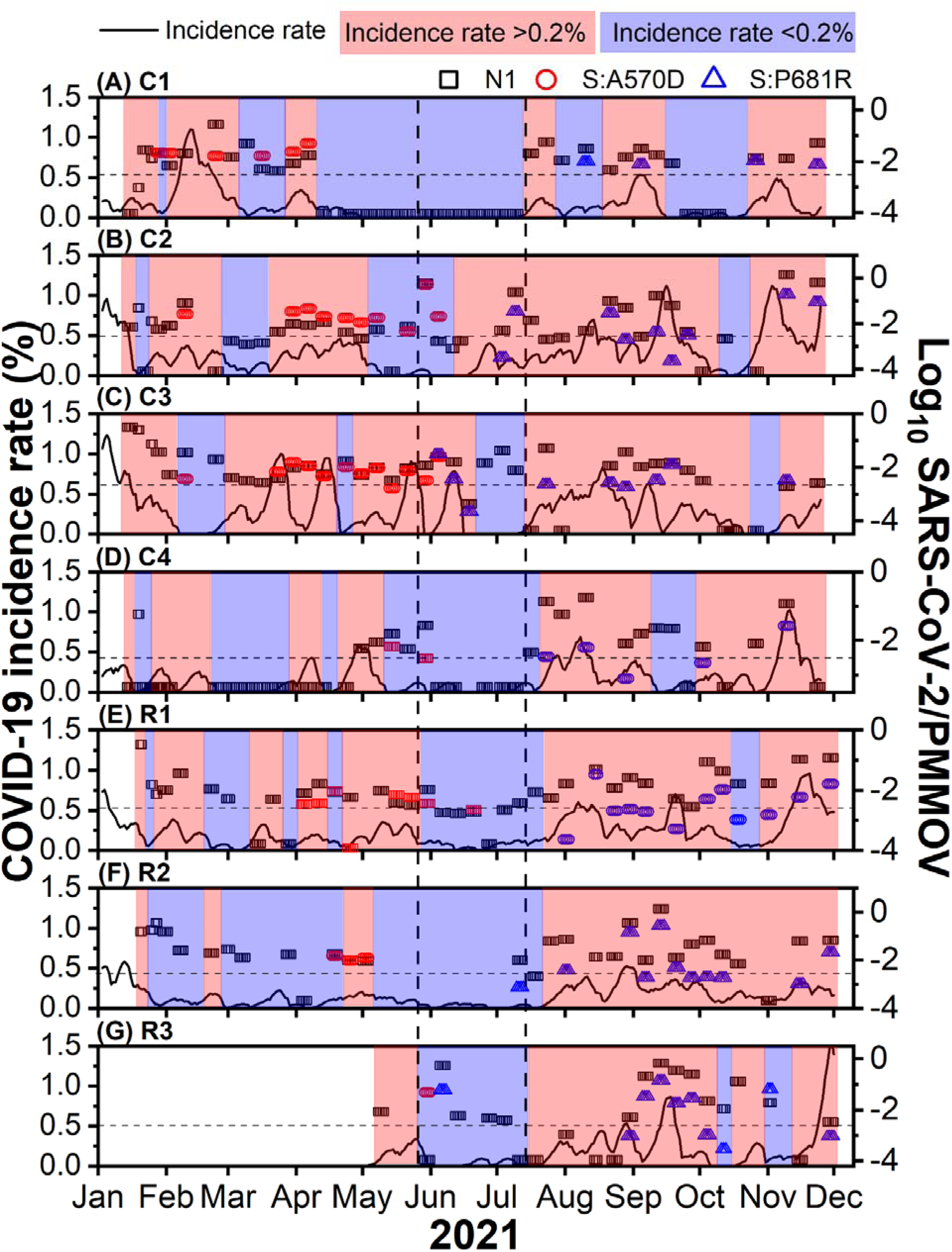
Comparisons between clinical epidemiology (COVID-19 incidence rate) and wastewater-based epidemiology (Log_10_ SARS-CoV-2/PMMOV). Open symbols represent WBE results (i.e., black rectangles represent the N gene, red circles represent the S:A570D mutation (potentially indicating the Alpha variant), and blue triangles represent S:P681R (potentially representing the Delta variant). The horizontal dashed lines show the threshold N/PMMOV of 10^-2.6^. We assigned about 10^-4^ of N/PMMOV to samples tested negative for the N gene to differentiate these samples from samples that were not measured. Black solid curves show the COVID-19 incidence rates. The monitoring period is separated either into the high (red) or the low level (blue) with the criteria of a 0.2% incidence rate. Vertical dashed lines indicate the monitoring period from May 28 to July 12 when the incidence rate on a county-scale was below 0.05%.

**Fig. 4.**
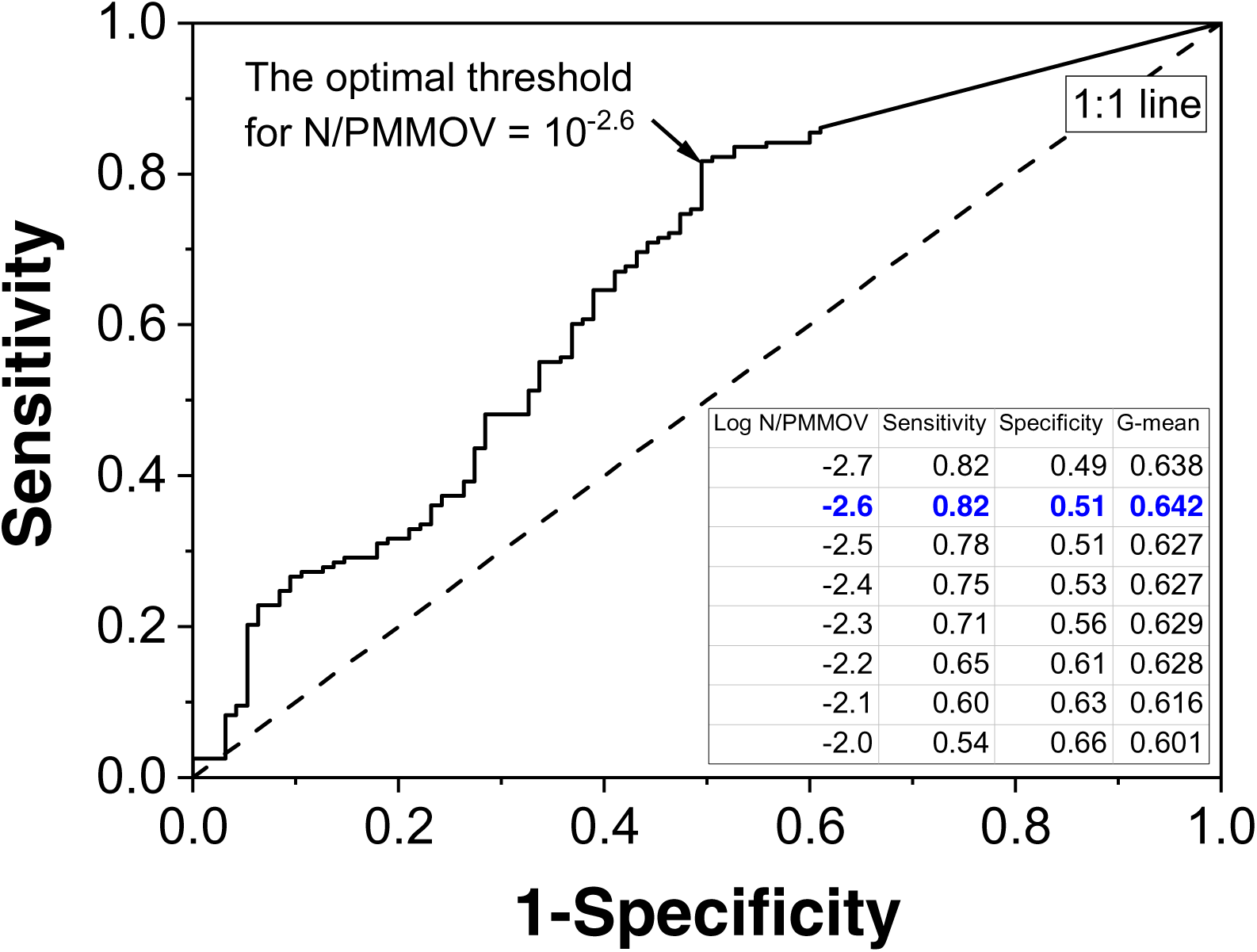
ROC curve with a series of N/PMMOV values when a COVID-19 incidence rate is 0.2%. The area under the curve (AUC) is 0.66 with 95% CI[0.59; 0.73]. The sensitivity and specificity with N/PMMOV of 10^-2.6^ are 0.82 and 0.51, respectively. The dashed line indicates a 1:1 line.

We determined a sensitivity of our WBE procedure, defined as the COVID-19 incidence rate at which viruses in sewage are detected at 50% probability. The data from C1 was used for a binary logistic regression analysis because the COVID-19 incidence rate is most reliable in C1 due to the mandatory COVID-19 testing. We found that the COVID-19 incidence rate is a reliable predictor to explain the probability of SARS-CoV-2 N gene detection (McFadden; p<0.001). Based on the logistic regression curve in **Fig. 5,** we determined 0.05% for the WBE sensitivity (or one infection case out of 2000 residents). **Fig. 6** shows the COVID-19 incidence rate in Champaign County, which includes all seven sewersheds during the same monitoring period as our WBE program. We found the incidence rate was less than the WBE sensitivity (0.05%) from May 27 to July 12 on a county-scale, suggesting that viruses would not have been detected from sewage if the WBE had been applied to the entire county during this low incidence rate period.

**Fig. 5.**
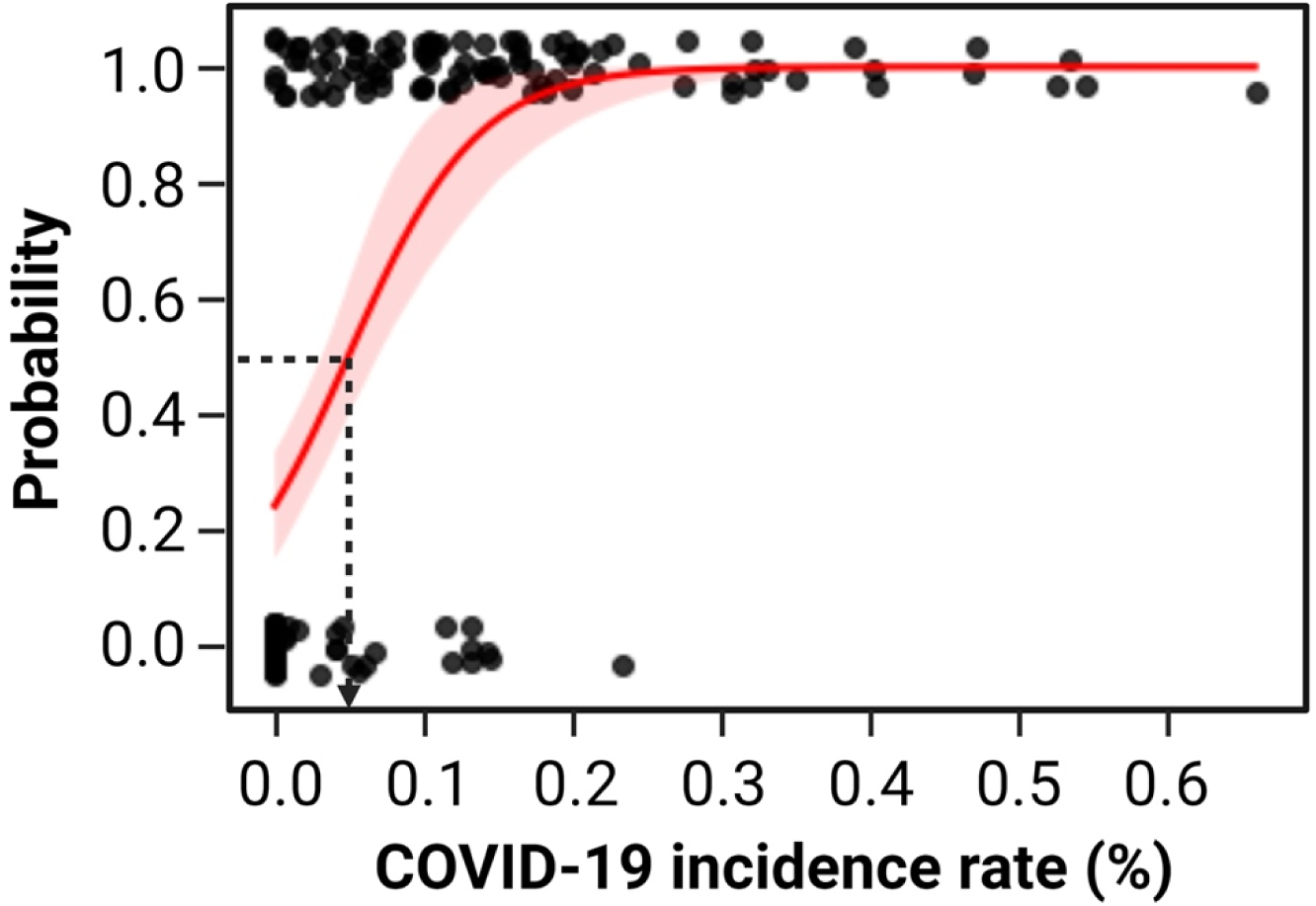
Probability of detecting SARS-CoV-2 N gene in sewage samples from location C1. Black closed-circles represent COVID-19 incidence rates on the x-axis. Either 1 or 0 was given to the y-value when sewage samples tested positive or negative for SARS-CoV-2, respectively. Logistic regression curves with a 95% confidence interval are represented by a red solid line and shade. McFadden’s Pseudo R-squared of the regression analysis was 0.34 (p<0.001), which is considered a good fit (McFadden, 2021). The Black dotted line indicates that the 0.05% incidence rate results in a 50% probability.

**Fig. 6.**
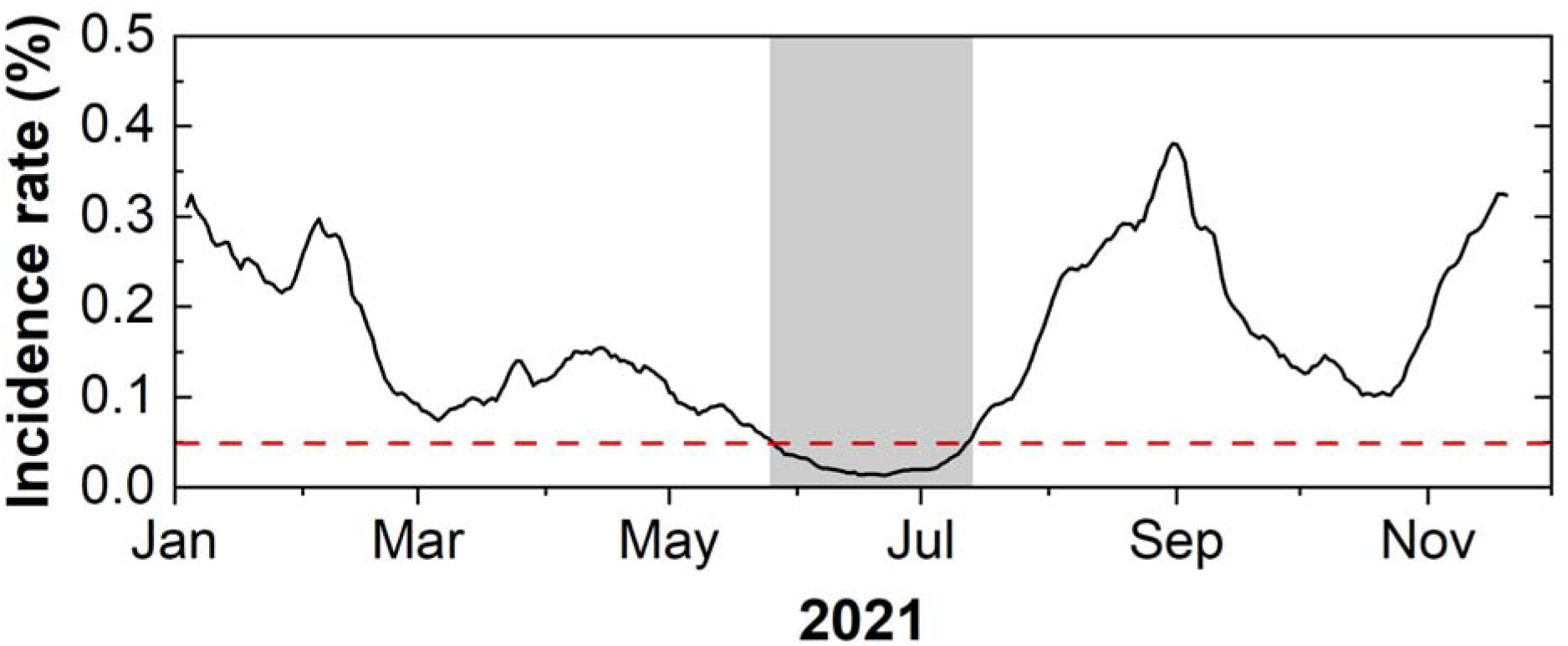
COVID-19 incidence rate in Champaign County. The red dashed line indicates a 0.05% of incidence rate with which viruses in sewage discharged from a county-scale sewershed are expected to be detected with 50% probability. The gray shade represents the monitoring period showing the incidence rates lower than 0.05%.

However, local outbreaks with incidence rates higher than 0.2% were reported from Locations C2 and C3 from May 27 to July 12 when the county-scale incidence rate was below the WBE sensitivity (0.05%). These examples show that incidence rates on neighborhood-scale sewersheds could be different from those on a county-scale sewershed. We found that the N/PMMOV values were higher than the threshold of 10^−2.6^ at Locations C2 and C3, which means the neighborhood-scale WBE correctly indicates these local outbreaks. More importantly, the introduction of the Delta variant to Champaign County (i.e., Locations C3 and R3) was first reported by our wastewater surveillance on June 6. The detection of the Delta variant by neighborhood scale WBE preceded the clinical epidemiology data. Three cases of Delta variant infections (GISAID ID: EPI_ISL_2885447, EPI_ISL_2885451, and EPI_ISL_2885452) were first confirmed by clinical tests on June 27 and reported on July 9 in Champaign County (**Fig. S8**). Also, these local outbreaks associated with the emerging variants would not have been identified if WBE was applied to county-scale sewershed because of the incidence rate below the WBE sensitivity.

To elucidate the effect of a sewershed size on SARS-CoV-2 N gene detection, we monitored sewage samples from another set of sewersheds, including three neighborhood schools (S1, S2, and S3) and one city-scale sewershed (City), from January to March 2022. This monitoring period presents a decreasing tendency of incidence rate after the outbreak caused by the Omicron variant. **Fig. 7A** shows WBE and clinical epidemiology data from city-scale sewershed. The incidence rate decreased steadily and became lower than 1% in February 2022. The detection rate of the N gene was 55% during the entire monitoring period (6 positives out of 11 samplings) but decreased to 29% (2 positives out of 7 samplings) in February 2022. The reduction of the N gene detection rate is expected due to the low incidence rate on a city-scale. The National Wastewater Surveillance System (NWSS) provides information on the detection rate of SARS-CoV-2 from a wastewater treatment plant in Champaign city (https://covid.cdc.gov/covid-data-tracker/#wastewater-surveillance; sewershed 655) (a blue dashed line in **Fig. 7A**). The detection rate by NWSS agrees with our wastewater surveillance program, showing a decreasing detection rate from February 2022. On the other hand, the average N gene detection rate from the three smaller-scale sewersheds was stable regardless of the city-scale incidence rate, showing a 64% of detection rate (21 positives out of 33 samplings) during the entire period and 62% during the low incidence rate from February 2022 (13 positives out of 21 samplings). Location B2 reported positives for N/PMMOV throughout the monitoring period with especially a high detection rate of 82% (6 positives out of 7 samplings) when the incidence rate on a city-scale was low (**Fig. 7B**). These findings corroborate that wastewater surveillance should be performed at a smaller-scale sewershed to discern local outbreaks when the COVID-19 incidence rate is low.

**Fig. 7.**
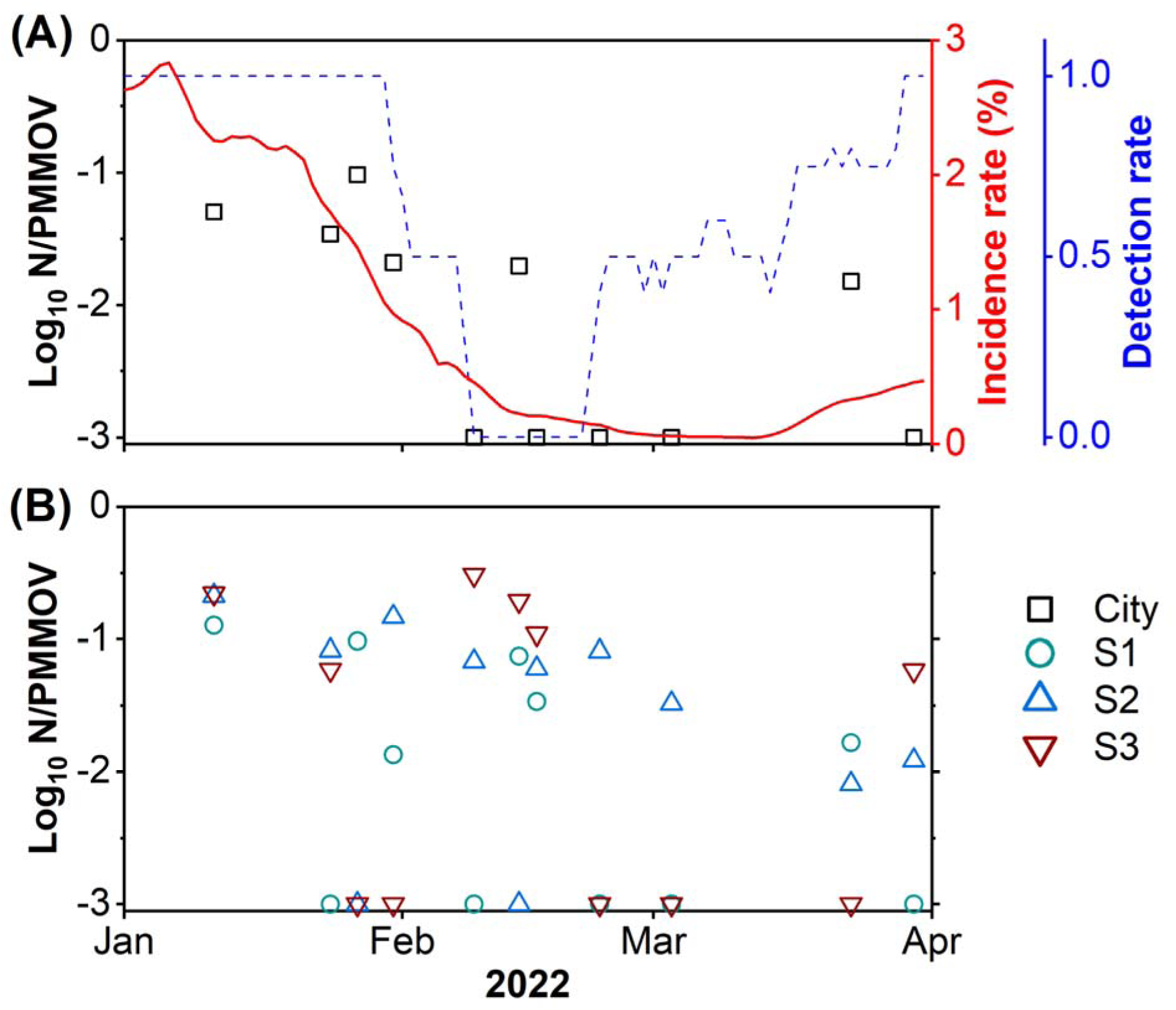
COVID-19 surveillance data at (A) one city-scale sewershed and (B) three neighborhood schools. A solid red line indicates the COVID-19 incidence rate from a city-scale sewershed. A blue dashed line represents the detection rate of the N gene published by the National Wastewater Surveillance System (NWSS).

### Implication

We suggested using a threshold of N/PMMOV to indicate a high level of the COVID-19 incidence rate. The ROC curve (**Fig. 4**) shows that sensitivity increased from 0.54 to 0.82 while the specificity decreased from 0.66 to 0.49 when N/PMMOV lowered from 10^-2.0^ to 10^-2.7^. Although we selected an unbiased threshold of 10^-2.6^ based on the highest geometric mean, different thresholds of N/PMMOV can be applied to the other sites considering regional characteristics such as local public health policies and the community’s acceptance of the COVID-19 risk (Dryhurst et al., 2020; Turska-Kawa and Pilch, 2022). Intensive care unit (ICU) occupancy rate, influenced by vulnerable populations and ICU bed capacity, may be one of the regional characteristics that should be considered to determine a threshold of N/PMMOV. If some regions had experienced a high ICU occupancy rate during the COVID-19 pandemic, COVID-19 put enormous stress on the local medical systems. Similar scenarios could happen again in the future. According to CDC, the maximum ICU occupancy rate significantly differs from site to site (http://covid.cdc.gov/covid-data-tracker). For example, in Illinois (USA), some counties, such as Cook County and Champaign County, have managed ICU beds well during the COVID-19 pandemic, showing 35% of maximum ICU bed occupancy. On the other hand, other counties, including Rock Island County (85%) and Effingham County (70%), experienced a more significant burden on ICU beds during the COVID-19 pandemic. Therefore, these counties may need to be more vigilant about the COVID-19 outbreak by setting a lower threshold.

Applying WBE to neighborhood-scale sewersheds may cost more than large-size sewersheds for monitoring the entire administrative area. To efficiently monitor multiple neighborhood-scale sewersheds, the WBE can be deployed to some representative places of diverse societal characteristics, such as socioeconomic status, demographics, or land use category, so that we can minimize the number of monitoring sites while obtaining information about the COVID-19 incidence representative of the entire area. The COVID-19 pandemic disproportionally has impacted people depending on demographic and socioeconomic status (Bassett et al., 2020; Krieger, 2020). Thus, those underserved regions could be chosen for wastewater surveillance. For example, we chose the nine sewersheds (Locations C2, C3, C4, R1, R2, R3, S1, S2, and S3) based on areas that reported fewer cases of COVID-19 testing than the average testing in Champaign County to attempt to supplement clinical testing data, while we selected another sewershed to represent the highly tested campus area (location C1) (**Table S8**). In addition, sample collection frequency can be optimized to operate WBE within a budget. Schoen et al. (2022) found that a sampling frequency of once every 4 days is not significantly different from daily sampling in estimating clinical epidemiology data. Therefore, we collected 4-day composite samples with a less frequent sampling frequency, once every 8.2 days.

We suggest that the low correlation coefficients between WBE and the clinical data can be explained by the fact that heterogeneity of clinical symptoms becomes significant with a low COVID-19 incidence rate in neighborhood-scale sewersheds. However, these correlation coefficients may become higher if our wastewater surveillance protocol is more sensitive to detecting the N gene from sewage samples. Although the median of our LODs was 3.78 log_10_ gc/L, which is within the reasonable range compared to other studies (3.0 log_10_ gc/L (10th percentile) to 6.1 log_10_ gc/L (90th percentile) (Pecson et al., 2021), the WBE sensitivity could be getting better as WBE is getting matured. Then, the correlation coefficients could be higher, and it could be possible to quantitatively estimate the COVID-19 incidence rate with N/PMMOV.

Our findings on the neighborhood-scale WBE have significant implications for public health. COVID-19 might be moving from a pandemic to an endemic phase (Veldhoen and Pedro Simas, 2021), which involves a difference in disease surveillance strategy. In an endemic phase, we may not be able to get as accurate statistics on COVID-19 by the clinical diagnosis as we did in the pandemic phase because people will be less likely to visit hospitals to get tested or medical support. This is because the illnesses may be self-limiting (Veldhoen and Pedro Simas, 2021). People also may have less accessibility to testing sites and use rapid COVID-19 test kits at home (Kost, 2022). In this context, WBE will play a pivotal role in the COVID-19 surveillance because WBE provides unbiased information about all residents in the sewershed. Lak et al. (2021) and Shi et al. (2021) discovered that local COVID-19 outbreaks could spread through the surrounding regions resulting in other outbreaks. Rader et al. (2020) explained the fast virus transmission to the surrounding areas with frequent contact among residents. These studies highlight the necessity of early detection of local COVID-19 outbreaks (Cariti et al., 2022; Wu et al., 2022a). Our study shows that WBE should be applied to neighborhood-scale sewershed to identify covert local outbreaks in advance and the introduction of emerging SARS-CoV-2 variants, which provides a warning for large-scale outbreaks.

## Conclusion

In this study, we applied WBE to seven neighborhood-scale sewersheds (an average catchment population was 1471) for eleven months. We found that WBE data from neighborhood-scale sewersheds were poorly correlated to clinical testing data when COVID-19 incidence was low. Thus, we suggest a threshold of N/PMMOV to indicate a high level of COVID-19 incidence rates, which can be assigned in collaboration with public health officials. We demonstrated that this approach could discern local COVID-19 outbreaks that a county-scale WBE would not detect due to the low disease incidence on a county scale. Our findings highlight that WBE should be applied to neighborhood-scale sewersheds when COVID-19 incidence is maintained at a low level.

## Data Availability

All data produced in the present work are contained in the manuscript

## Acknowledgment

We acknowledge the funding from the Grainger College of Engineering, the JUMP-ARCHES program of OSF Healthcare in conjunction with the University of Illinois, and the VinUni Illinois Smart Health Center. We thank Brad Bennett and Bruce Rabe at the Urbana-Champaign Sanitary District and Haley Turner and Travis Ramme at the Rantoul Wastewater Treatment Plant for providing us with influent wastewater. The authors also acknowledge Kip Stevenson for sampling deployment, and Yuqing Mao, Matthew Robert Loula, Aashna Patra, Kristin Joy Anderson, Mikayla Diedrick, Hubert Lyu, Hamza Elmahi Mohamed, Jad R Karajeh, Runsen Ning, Rui Fu, and Kyukyoung Kim for sewage sampling and processing. We also acknowledge Dr. Awais Vaid for guidance on sampling site selection.

