## Supplemental Figures and Tables for "Application of neighborhood-scale wastewater-based epidemiology in low COVID-19 incidence situations"

**Eight figures and eight tables are included.**

**Table S1.** The minimum recommended meta-information on sewage samples^1)^

| Location name | C1 | C2 | C3 | C4 | R1 | R2 | R3 | S1 | S2 | S3 | City |
| --- | --- | --- | --- | --- | --- | --- | --- | --- | --- | --- | --- |
| Sample location type | Street line manhole | | | | | | | Sewer line before septic tank | | | Influent to wastewater treatment plant |
| Population served | 1675 | 1260 | 853 | 859 | 2402 | 2160 | 1088 | 9094 | 3547 | 5547 | 144097 |
| Combined or separated system | Separated | | | | | | | | | | |
| Sample collection type | Time-weighted composite samples | | | | | | | | | | |
| Sample matrix | Sludge from raw wastewater | | | | | | | | | | |
| Sample date | January 14, 2021 - November 25, 2021 | | | | January 20, 2021 - December 2, 2021 | | May 8, 2021 to Dec 2, 2021 | January 11, 2022 - March 30, 2022 | | | |
| Sample time | Composite samples were delivered between 8 am to 12 pm to the laboratory. | | | | | | | | | | |
| Sample location | Champaign | Urbana | Champaign | Champaign | Rantoul | | | Rural area of Champaign County | | | Urbana |
| Pre-concentration storage temperature | Samples were delivered to the laboratory on ice and processed without freezing on the same day. | | | | | | | | | | |
| Concentration method and citation | RNA extraction from sludge^2)^ | | | | | | | | | | |
| Recovery control name & efficiency | BCoV was used for recovery control and the recovery efficiencies are summarized in Fig. S5. | | | | | | | | | | |
| Extraction method & citation | Viral RNA Mini Kit (Quigen, German) | | | | | | | | | | |
| Amount of sample processed | Approximately 1 or 2 L | | | | | | | | | | |
| Extraction blanks results | Wastewater discharged from a food processing plant was treated together with sewage samples. These controls tested negative for the SARS-CoV-2 N gene. | | | | | | | Extraction blanks were not included in these data. | | | |
| PCR type | RT-qPCR | | | | | | | | | | |
| SARS-CoV-2 concentrations | Between below limit of detection to 9.2$\times$10^6^ (gc/L) | | | | | | | | | | |
| Target gene | N1 gene (CDC) | | | | | | | | | | |
| Endogenous wastewater control name & concentration | PMMoV was used as an internal control and the concentrations are summarized in Fig. S4 | | | | | | | | | | |
| Required MIQE guideline | Summarized in Table S3 | | | | | | | | | | |

1) McClary-Gutierrez et al. (2021)
2) Wolfe et al. (2021)

**Table S2.** Summary of six RT-qPCR assays

| Target species | Target gene or mutation | Primer name | Sequence (5’-3’) | GC content (%) | Tm (℃) | Amplicon size (bp) | Purpose |
| --- | --- | --- | --- | --- | --- | --- | --- |
| SARS-CoV-2 | N^1)^ | CDC_N1_Forward | GACCCCAAAATCAGCGAAAT | 45.0 | 61.1 | 72 | Total SARS-CoV-2 |
|  |  | CDC_N1_Reverse | TCTGGTTACTGCCAGTTGAATCTG | 45.8 | 64.5 |  |  |
|  |  | CDC_N1_Probe | (FAM)ACCCCGCATTACGTTTGGTGGACC(IBFQ) | 58.3 | 70.3 |  |  |
|  | S:A570D^2)^ | Alpha_Forward | ACAATTTGGCAGAGACATCGA | 42.9 | 62.3 | 85 | Alpha variant |
|  |  | Alpha_Reverse | AGAACATGGTGTAATGTCAAGAATC | 36.0 | 61.7 |  |  |
|  |  | Alpha_Probe | (HEX)ACTGATGCTGTCCGTGATCCACAG(IBFQ) | 54.2 | 67.8 |  |  |
|  | S:P681R^3)^ | Delta_Forward | ATCAGACTCAGACTAATTCACG | 40.9 | 59.6 | 87 | Delta variant |
|  |  | Delta_Reverse | TTTCTGCACCAAGTGACATA | 40.0 | 59.7 |  |  |
|  |  | Delta_Probe | (FAM)CGGGCACGTAGTGTAGCTAGTCAA(IBFQ) | 54.2 | 67.2 |  |  |
| PMMOV^4)^ | | PMMOV_Forward | ATGAGAGTGGTTTGACCTTAAC | 40.9 | 60.4 | 95 | Normalization to human feces |
|  |  | PMMOV_Reverse | CGAACCTTCCTCCTTTGATG | 50.0 | 60.3 |  |  |
|  |  | PMMOV_Probe | (HEX)AGGCCTACCGAAGCAAATGTCGCA(IBFQ) | 54.2 | 70.1 |  |  |
| TV^5)^ | | TV_Forward | GTGCGCATCCTTGAGACAAT | 50.0 | 63.0 | 133 | PCR inhibition test |
|  |  | TV_Reverse | TTGGAGCCGGGTAGAAACAT | 50.0 | 63.5 |  |  |
|  |  | TV_Probe | (FAM)CCCTTGGAAACCTCACCAGGAATCA(IBFQ) | 52.0 | 67.9 |  |  |
| BCoV^6)^ | | BCoV_Forward | CTAGTAACCAGGCTGATGTCAATACC | 46.2 | 64.2 | 89 | Recovery efficiency |
|  |  | BCoV_Reverse | GGCGGAAACCTAGTCGGAATA | 52.4 | 63.5 |  |  |

1) This assay was designed by U.S. CDC (2020). Twist Synthetic SARS-CoV-2 RNA Control 2 (102024, TWIST Bioscience, CA, USA) was used as a standard sample.
2) This assay was designed by Lee et al. (2021). Twist Synthetic SARS-CoV-2 RNA Control 14 (103907, TWIST Bioscience, CA, USA) was used as a standard sample.
3) Twist Synthetic SARS-CoV-2 RNA Control 23 (104533, TWIST Bioscience, CA, USA) was used as a standard sample.
4) Coding sequences for replicase protein were targeted (Genbank: MN496154.1). Synthetic DNA was designed by Integrated DNA Technologies (IDT, IA, USA). The sequence is: 5’-TACAATGCTTTGTCAGAAATCTCAATTCTTAAAGACAGTGACAAGTTTGATGTTGATGTTTTTTCCCGGATGTGTAATACATTAGGCGTAGATCCATTGGTGGCAGCAAAGGTAATGGTAGCTGTGGTTTCAAATGAGAGTGGTTTGACCTTAACGTTTGAGAGGCCTACCGAAGCAAATGTCGCACTTGCATTGCAACCGACAATTACATCAAAGGAGGAAGGTTCGTTGAAGATTGTGTCGTCAGACGTAGGTGAGTCCTCAATCAAGGAAGTGGTTCGAAAATCAGAGATTTCTATGCTTGGTCTAACAGGCAACACAGTGTCCGATGAGTTCCAAAGAAGTACAGAAATCGAGTCG-3’
5) This assay was designed by Fuzawa et al. (2020). FLA45_gp1 gene is targeted (Genbank: NC_043512.1). The sequence of standard sample for the NSP1 gene of TV (Integrated DNA Technologies, USA) is: 5’-AGAATTGGACCGAATTTGGCACACACTCAGAATTTGGTGTGCGCATCCTTGAGACAATAACAGGCACAATACCCCCTTGGAAACCTCACCAGGAATCAATATCTGAAGTTCTGGACGACCTCACACACGGTAAAGTCCAAACAGGTGATGATGTTTCTACCCGGCTCCAAAGGTTGAGCGACACTATCAAAGATCTGAGTGTCATGGCTTGTGATCCCTCTGCACCGCCCGAAGTTGCGC-3’
6) This assay was designed by Cho et al. (2013). N gene is targeted (Genbank: LC494177.1). Synthetic DNA was designed by Integrated DNA Technologies (IDT, IA, USA). The sequence is: 5’-AGCTAAAGGGTACTGGTACAGACACAACAGACGTTCCTTTAAAACAGCCGATGGCAACCAGCGTCAATTGCTTCCACGATGGTATTTTTACTATCTTGGAACAGGACCGCATGCCAAAGACCAGTATGGCACCGACATTGACGGAGTCTTCTGGGTCGCTAGTAACCAGGCTGATGTCAATACCCCGGCTGACATTCTCGATCGGGACCCAAGTAGCGATGAGGCTATTCCGACTAGGTTTCCGCCTGGCACGGTACTCCCTCAGGGTTACTATATTGAAGGCTCAGGAAGGTCTGCTCCTAATTCCAGATCTACTTCACGCGCATCCAGTAGAGCCTCTAGTGCAGGATCGCGCAGTAG-3’
7) Primer pair specificity was checked by the primer-blast tool (National Center of Biotechnology Information). Each pair of primers were blasted with Homo sapiens (txid:9606). We confirmed that our primers do not target any sequences of human genes.

**Table S3.** The checklist from MIQE guidelines and relevant information for this study

| **Item to check** | **Location** |
| --- | --- |
| 1. Experimental design | |
| Definition of experimental and control groups | Throughout Materials and Methods section |
| Number within each group | Materials and Methods (Analysis of viral genomes) |
| 2. Sample | |
| Description | Materials and Methods (Collection of raw sewage composite samples) |
| Volume/mass of sample processed | Materials and Methods (Collection of raw sewage composite samples) |
| Processing procedure | Materials and Methods (Sample processing and viral nucleic acid extraction) |
| Sample storage conditions and duration | RNA samples were stored at -80 ℃ for less than 1 year |
| 3. Nucleic acid extraction | |
| Procedure and/or instrumentation | Materials and Methods (Analysis of viral genomes) |
| Name of kit and details of any modifications | Materials and Methods (Analysis of viral genomes) |
| Contamination assessment (DNA or RNA) | Materials and Methods (Analysis of viral genomes) |
| Nucleic acid quantification | Materials and Methods (Analysis of viral genomes) |
| Instrument and method | Materials and Methods (Analysis of viral genomes) |
| Inhibition testing (C_q_ dilutions, spike, or other) | Materials and Methods (Fig. S6) |
| 4. Reverse transcription | |
| Complete reaction conditions | Materials and Methods (Analysis of viral genomes) |
| Amount of RNA and reaction volume | Materials and Methods (Analysis of viral genomes) |
| Reverse transcriptase and concentration | Materials and Methods (Analysis of viral genomes) |
| Temperature and time | Materials and Methods (Analysis of viral genomes) |
| Manufacturer of reagents and catalog numbers | Materials and Methods (Analysis of viral genomes) |
| 5. qPCR target information | |
| Gene symbol | Materials and Methods, Table S2 |
| Sequence accession number | Materials and Methods, Table S2 |
| Location of amplicon | Materials and Methods, Table S2 |
| Amplicon length | Materials and Methods, Table S2 |
| In silico specificity screen (BLAST, and so on) | Materials and Methods, Table S2 |
| 6. qPCR oligonucleotides | |
| Primer sequences | Materials and Methods, Table S2 |
| Manufacturer of oligonucleotides | Materials and Methods, Table S2 |
| 7. qPCR protocol | |
| Complete reaction conditions | Materials and Methods (Analysis of viral genomes) |
| Reaction volume and amount of DNA | Materials and Methods (Analysis of viral genomes) |
| Primer (probe) concentrations | Materials and Methods (Analysis of viral genomes) |
| Polymerase identity and concentration | Materials and Methods (Analysis of viral genomes) |
| Buffer/kit identity and manufacturer | Materials and Methods (Analysis of viral genomes) |
| Manufacturer of plates/tubes and catalog number | Materials and Methods (Analysis of viral genomes) |
| Complete thermocycling parameters | Materials and Methods (Analysis of viral genomes) |
| Manufacturer of qPCR instrument | Materials and Methods (Analysis of viral genomes) |
| 8. qPCR validation | |
| Specificity (gel, sequence, melt, or digest) | Materials and Methods (Analysis of viral genomes) |
| For SYBR Green I, C_q_ of the NTC | Materials and Methods (Analysis of viral genomes) |
| Calibration curves with slope and *y* intercept | Materials and Methods, Fig. S7 |
| PCR efficiency calculated from slope | Materials and Methods, Fig. S7 |
| *r*2 of calibration curve | Materials and Methods, Fig. S7 |
| Linear dynamic range | Materials and Methods, Fig. S7 |
| C_q_ variation at LOD | Materials and Methods, Table S4 |
| Evidence for LOD | Materials and Methods, Table S4 |
| 9. Data analysis | |
| qPCR analysis program (source, version) | Materials and Methods (Analysis of viral genomes) |
| Method of C_q_ determination | Materials and Methods (Analysis of viral genomes) |
| Outlier identification and disposition | Materials and Methods (Analysis of viral genomes) |
| Results for NTCs | Materials and Methods (Analysis of viral genomes) |
| Description of normalization method | Materials and Methods (Analysis of viral genomes) |
| Number and concordance of biological replicates | Materials and Methods (Analysis of viral genomes) |
| Number and stage of technical replicates | Materials and Methods (Analysis of viral genomes) |
| Repeatability (intra assay variation) | Materials and Methods (Analysis of viral genomes) |
| Statistical methods for results significance | Each figure |
| Software (source, version) | Materials and Methods (Analysis of viral genomes) |
| Data transparency | Raw data available upon request |

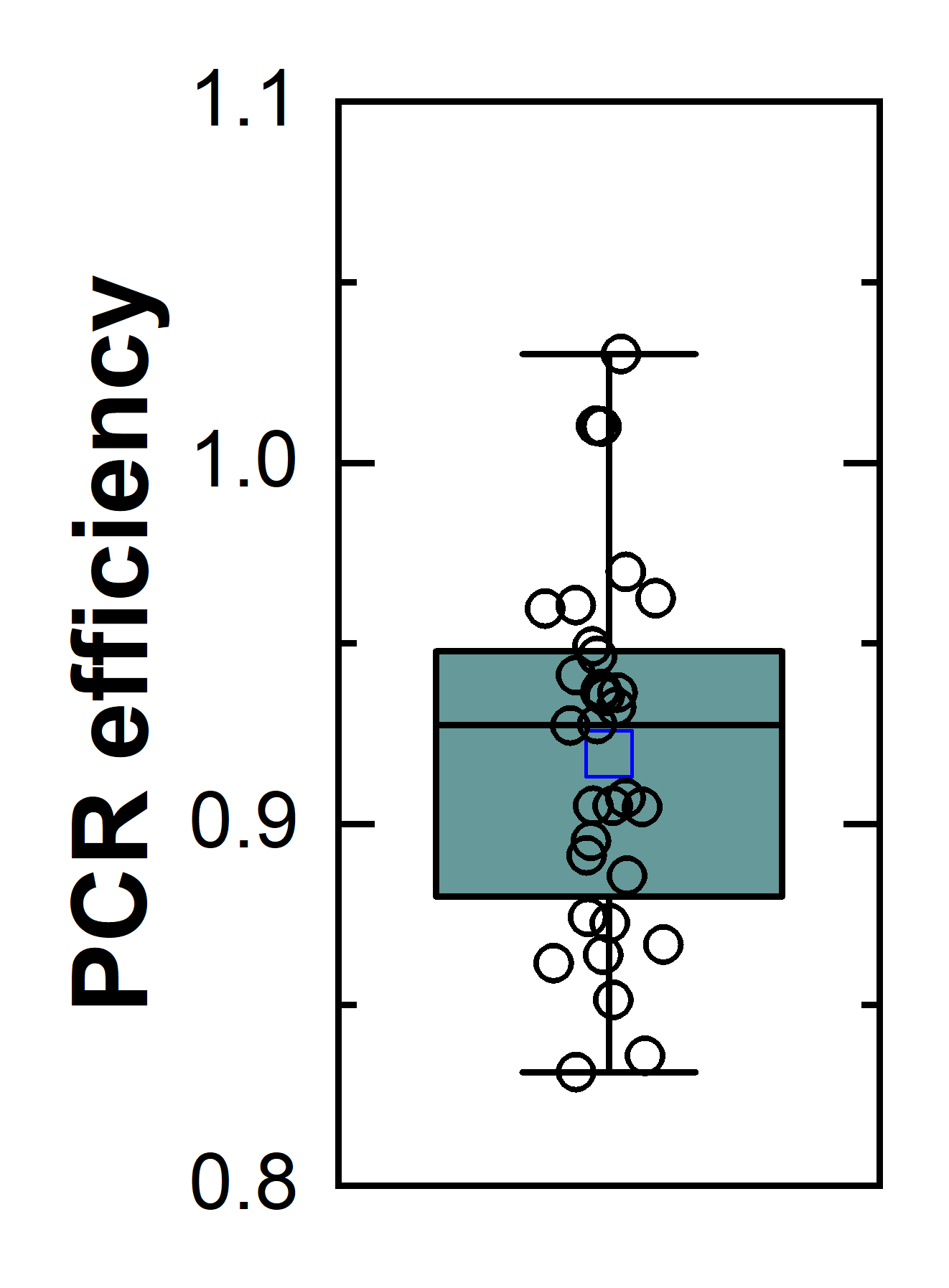

**Fig. S1.** PCR efficiency of Taqman-based RT-qPCR assays for the N gene, S:A570D, and S:P681R mutation.

**Table S4.** Summary of limit of detection (LOD) and limit of quantification (LOQ) for RT-qPCR assays

| Target | LOD | | LOQ | |
| --- | --- | --- | --- | --- |
|  | gc/$\mu$L | gc/reaction | gc/$\mu$L | gc/reaction |
| SASRS-CoV-2 N | 0.44 | 2.2 | 10.0 | 50.0 |
| S:A570D mutation | 0.44 | 2.2 | 10.0 | 50.0 |
| S:P681R mutation | 0.44 | 2.2 | 10.0 | 50.0 |
| PMMOV | 1.60 | 8.0 | 12.6 | 62.9 |
| TV | 0.83 | 4.2 | 6.3 | 31.5 |

**Table S5**. Limit of detection (LOD) of N gene concentration of sewage samples (gc/L)

| Date | C1 | C2 | C3 | C4 | Date | R1 | R2 | R3 |
| --- | --- | --- | --- | --- | --- | --- | --- | --- |
| 1/14/21 | 1,998 | 4,484 | 16,923 | 3,486 | 1/20/21 | 10,164 | 14,141 | - |
| 1/19/21 | 13,518 | 79,279 | 10,179 | 68,723 | 1/25/21 | 5,377 | 1,000,000 | - |
| 1/21/21 | 4,250 | 29,820 | - | - | 1/27/21 | 1,040 | 11,855 | - |
| 1/25/21 | 9,664 | 15,745 | 71,895 | 15,985 | 1/30/21 | 1,581 | 15,155 | - |
| 1/27/21 | 16,358 | 34,320 | 2,581 | 45,286 | 2/6/21 | 4,237 | 60,307 | - |
| 2/1/21 | 6,028 | 11,573 | 29,804 | 15,378 | 2/20/21 | 6,723 | 176,367 | - |
| 2/8/21 | 7,217 | 4,740 | 2,771 | 13,518 | 2/28/21 | 786 | 3,779 | - |
| 2/22/21 | 817 | 1,345 | 939 | 3,811 | 3/6/21 | - | 82,090 | - |
| 3/1/21 | 694 | 2,407 | 1,886 | 3,278 | 3/13/21 | 4,691 | - | - |
| 3/8/21 | 3,927 | 28,490 | 2,655 | 10,681 | 3/20/21 | 3,884 | - | - |
| 3/15/21 | 4,052 | 25,463 | 20,721 | 8,605 | 3/27/21 | 17,316 | 23,109 | - |
| 3/22/21 | 1,729 | 29,750 | 1,178 | 9,589 | 4/3/21 | 5,023 | 167,619 | - |
| 3/29/21 | 2,195 | 90,956 | 30,352 | 6,209 | 4/10/21 | 5,678 | - | - |
| 4/5/21 | 3,206 | 181,818 | 1,406 | 6,851 | 4/17/21 | 1,278 | 6,847 | - |
| 4/12/21 | 2,509 | 167,619 | 6,336 | 9,020 | 4/24/21 | 43,286 | 144,156 | - |
| 4/22/21 | 2,415 | 6,017 | 3,953 | 17,298 | 5/1/21 | 58,761 | - | - |
| 4/29/21 | 5,892 | 63,907 | 5,210 | 6,219 | 5/8/21 | 12,821 | - | 45,432 |
| 5/6/21 | 1,371 | 43,409 | 4,790 | 52,646 | 5/15/21 | 20,244 | - | - |
| 5/13/21 | 961 | 10,730 | 1,377 | 1,606 | 5/22/21 | 18,481 | - | - |
| 5/20/21 | 1,658 | 789 | 5,333 | 10,531 | 5/29/21 | 2,143 | - | 920 |
| 5/28/21 | 1,594 | 532 | 1,183 | 8,205 | 6/5/21 | 4,061 | - | 3,970 |
| 6/3/21 | 1,532 | 2,374 | 2,907 | 34,929 | 6/12/21 | 14,286 | - | 2,916 |
| 6/10/21 | 854 | 82,051 | 561 | 19,919 | 6/19/21 | 256 | - | - |
| 6/13/21 | 71 | 50 | 503 | - | 6/26/21 | 7,576 | - | 1,773 |
| 6/24/21 | 1,871 | - | 20,786 | 21,989 | 7/3/21 | 1,912 | - | 5,724 |
| 7/2/21 | 1,985 | 8,590 | 12,546 | 4,303 | 7/10/21 | 2,783 | 984 | 2,754 |
| 7/8/21 | 2,082 | 962 | 10,256 | 8,690 | 7/17/21 | 2,267 | 8,903 | 33,951 |
| 7/15/21 | 1,590 | 758 | 664 | 11,126 | 7/24/21 | 177,419 | 1,128,205 | - |
| 7/22/21 | 8,121 | 1,102 | 8,690 | 3,999 | 7/31/21 | 2,146 | 14,981 | 2,105 |
| 7/29/21 | 1,586 | 860 | 7,381 | 4,916 | 8/14/21 | 5,864 | 4,405 | 41,509 |
| 8/9/21 | 2,127 | 638 | - | 6,947 | 8/22/21 | 3,654 | 8,366 | 47,930 |
| 8/20/21 | 1,686 | 14,712 | 3,480 | - | 8/29/21 | 8,691 | 11,642 | 31,977 |
| 8/27/21 | 1,255 | 3,001 | 5,299 | 1,751 | 9/5/21 | 10,480 | 15,490 | 4,388 |
| 9/3/21 | 4,603 | 2,097 | 120,104 | 11,767 | 9/12/21 | - | 2,907 | 5,493 |
| 9/10/21 | 2,717 | 1,813 | 14,747 | 18,861 | 9/19/21 | 3,030 | 11,372 | 7,189 |
| 9/17/21 | 9,427 | 50,271 | 16,494 | 35,285 | 9/26/21 | 20,530 | 4,549 | 6,874 |
| 9/24/21 | 1,910 | 4,930 | 6,209 | 75,485 | 10/3/21 | 12,857 | 3,194 | 7,256 |
| 10/1/21 | 3,714 | 7,588 | 31,283 | 4,346 | 10/10/21 | 4,991 | 3,101 | 2,573 |
| 10/11/21 | 2,504 | 2,137 | 3,161 | 46,049 | 10/17/21 | 4,666 | 3,138 | 3,898 |
| 10/25/21 | 1,751 | 1,267 | 25,463 | 7,143 | 10/31/21 | 3,602 | 4,968 | 84,034 |
| 11/8/21 | 6,587 | 4,285 | 9,012 | 6,450 | 11/14/21 | 14,913 | 8,326 | 8,787 |
| 11/22/21 | 45,380 | 81,580 | 29,993 | 265,460 | 11/28/21 | 7,376 | 15,842 | 25,985 |

**Table S6**. Limit of quantification (LOQ) of N gene concentration of sewage samples (gc/L)

| Date | C1 | C2 | C3 | C4 | Date | R1 | R2 | R3 |
| --- | --- | --- | --- | --- | --- | --- | --- | --- |
| 1/14/21 | 45,413 | 101,911 | 384,615 | 79,224 | 1/20/21 | 231,000 | 321,388 | - |
| 1/19/21 | 307,220 | 1,801,802 | 231,348 | 1,561,890 | 1/25/21 | 122,205 | 22,727,273 | - |
| 1/21/21 | 96,590 | 677,736 | - | - | 1/27/21 | 23,632 | 269,432 | - |
| 1/25/21 | 219,635 | 357,846 | 1,633,987 | 363,306 | 1/30/21 | 35,939 | 344,442 | - |
| 1/27/21 | 371,782 | 780,001 | 58,663 | 1,029,230 | 2/6/21 | 96,302 | 1,370,614 | - |
| 2/1/21 | 137,009 | 263,016 | 677,369 | 349,498 | 2/20/21 | 152,788 | 4,008,337 | - |
| 2/8/21 | 164,029 | 107,721 | 62,988 | 307,220 | 2/28/21 | 17,862 | 85,881 | - |
| 2/22/21 | 18,570 | 30,570 | 21,347 | 86,624 | 3/6/21 | - | 1,865,672 | - |
| 3/1/21 | 15,784 | 54,699 | 42,866 | 74,491 | 3/13/21 | 106,610 | - | - |
| 3/8/21 | 89,258 | 647,501 | 60,337 | 242,748 | 3/20/21 | 88,282 | - | - |
| 3/15/21 | 92,092 | 578,704 | 470,921 | 195,557 | 3/27/21 | 393,546 | 525,210 | - |
| 3/22/21 | 39,303 | 676,133 | 26,780 | 217,929 | 4/3/21 | 114,168 | 3,809,524 | - |
| 3/29/21 | 49,875 | 2,067,183 | 689,810 | 141,117 | 4/10/21 | 129,049 | - | - |
| 4/5/21 | 72,860 | 4,132,231 | 31,944 | 155,715 | 4/17/21 | 29,049 | 155,618 | - |
| 4/12/21 | 57,033 | 3,809,524 | 144,004 | 204,997 | 4/24/21 | 983,768 | 3,276,272 | - |
| 4/22/21 | 54,887 | 136,752 | 89,839 | 393,140 | 5/1/21 | - | 1,335,470 | - |
| 4/29/21 | 133,906 | 1,452,433 | 118,399 | 141,339 | 5/8/21 | 291,375 | - | 1,032,551 |
| 5/6/21 | 31,150 | 986,558 | 108,863 | 1,196,494 | 5/15/21 | 460,087 | - | - |
| 5/13/21 | 21,830 | 243,867 | 31,305 | 36,499 | 5/22/21 | 420,027 | - | - |
| 5/20/21 | 37,688 | 17,938 | 121,194 | 239,338 | 5/29/21 | 48,715 | - | 20,899 |
| 5/28/21 | 36,226 | 12,098 | 26,876 | 186,480 | 6/5/21 | 92,285 | - | 90,219 |
| 6/3/21 | 34,807 | 53,958 | 66,076 | 793,840 | 6/12/21 | 324,675 | - | 66,279 |
| 6/10/21 | 19,406 | 1,864,802 | 12,747 | 452,714 | 6/19/21 | 5,823 | - | - |
| 6/13/21 | 1,612 | 1,136 | 11,427 | - | 6/26/21 | 172,191 | - | 40,290 |
| 6/24/21 | 42,527 | - | 472,411 | 499,750 | 7/3/21 | 43,459 | - | 130,081 |
| 7/2/21 | 45,123 | 195,236 | 285,144 | 97,804 | 7/10/21 | 63,243 | 22,375 | 62,588 |
| 7/8/21 | 47,312 | 21,869 | 233,100 | 197,492 | 7/17/21 | 51,517 | 202,351 | 771,605 |
| 7/15/21 | 36,145 | 17,223 | 15,093 | 252,864 | 7/24/21 | 4,032,258 | 25,641,026 | - |
| 7/22/21 | 184,570 | 25,057 | 197,502 | 90,876 | 7/31/21 | 48,771 | 340,483 | 47,840 |
| 7/29/21 | 36,047 | 19,553 | 167,740 | 111,726 | 8/14/21 | 133,262 | 100,120 | 943,396 |
| 8/9/21 | 48,344 | 14,499 | 79,083 | 157,878 | 8/22/21 | 83,056 | 190,132 | 1,089,325 |
| 8/20/21 | 38,314 | 334,353 | 120,438 | - | 8/29/21 | 197,531 | 264,582 | 726,744 |
| 8/27/21 | 28,526 | 68,201 | 2,729,630 | 39,806 | 9/5/21 | 238,180 | 352,051 | 99,721 |
| 9/3/21 | 104,610 | 47,650 | 335,149 | 267,437 | 9/12/21 | - | 66,079 | 124,844 |
| 9/10/21 | 61,759 | 41,196 | 374,869 | 428,669 | 9/19/21 | 68,871 | 258,448 | 163,393 |
| 9/17/21 | 214,259 | 1,142,531 | 141,115 | 801,925 | 9/26/21 | 466,592 | 103,385 | 156,234 |
| 9/24/21 | 43,418 | 112,039 | 710,985 | 1,715,560 | 10/3/21 | 292,205 | 72,595 | 164,914 |
| 10/1/21 | 84,401 | 172,449 | 71,842 | 98,765 | 10/10/21 | 113,422 | 70,472 | 58,480 |
| 10/11/21 | 56,899 | 48,577 | 578,704 | 1,046,572 | 10/17/21 | 106,052 | 71,317 | 88,588 |
| 10/25/21 | 39,788 | 28,787 | 204,813 | 162,339 | 10/31/21 | 81,865 | 112,918 | 1,909,855 |
| 11/8/21 | 149,701 | 97,398 | 681,663 | 146,601 | 11/14/21 | 338,941 | 189,233 | 199,696 |
| 11/22/21 | 1,031,353 | 1,854,084 | 527,426 | 6,033,183 | 11/28/21 | 167,631 | 360,036 | 590,563 |

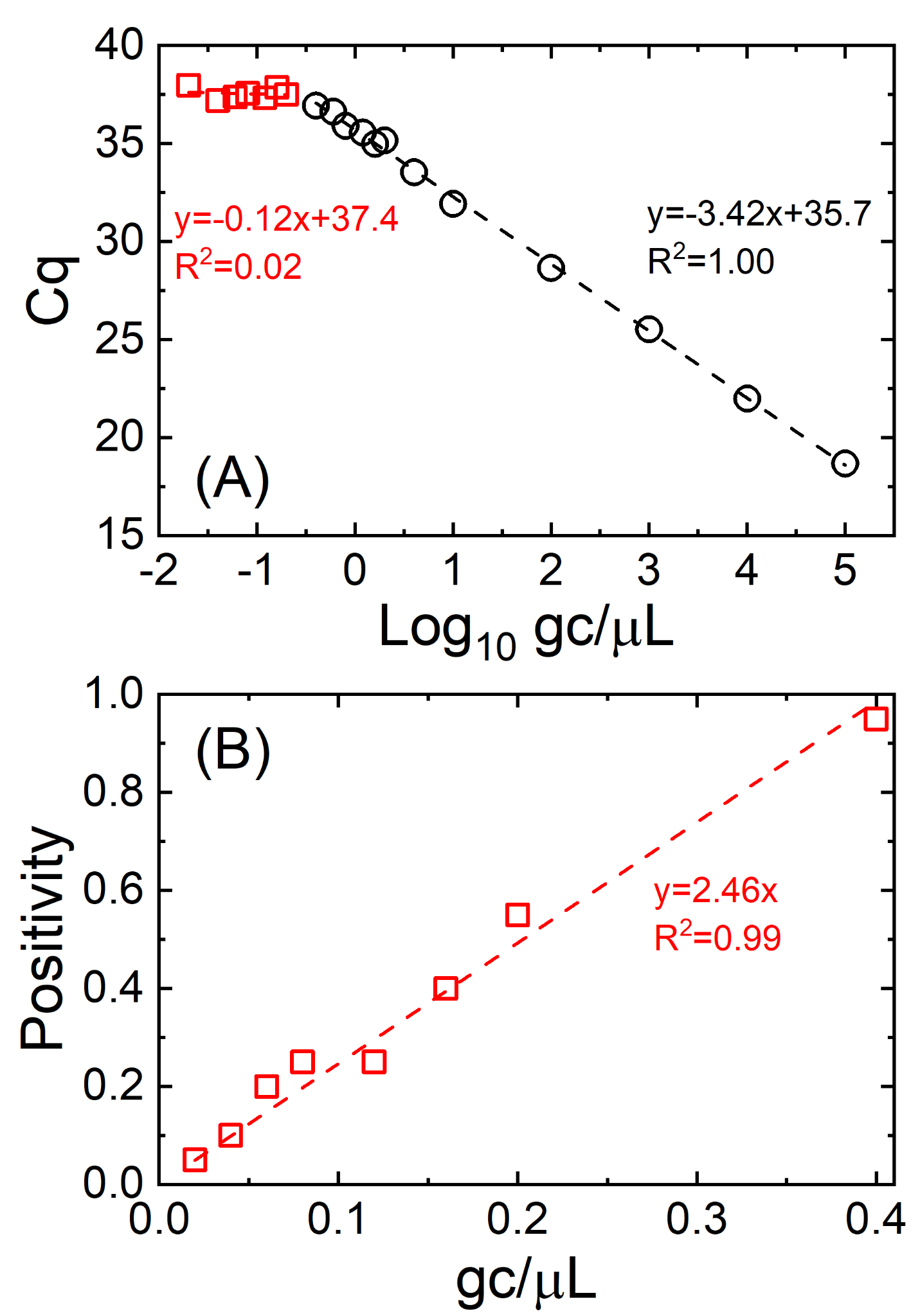

**Fig. S2.** Cq values and positivity of serial dilutions of synthetic RNA controls. (A) Average Cq values of 20 replicates, excluding undetermined Cq values. Black circles indicate samples whose technical replicates do not show undetermined Cq while red rectangles represent samples including at least one replicate showing undetermined Cq values. When the RNA concentrations were less than 0.2 gc/$\mu$L (or 1 gc/reaction), the Cq values, which were calculated by averaging positive Cq values replicates while excluding the undetermined Cq, did not follow the linear relationship (ANOVA; p>0.05) (B) Positivity of samples with one or more undetermined Cq values. The positivity shows a good correlation with the true RNA concentration ranging from 0 to 0.4 gc/$\mu$L when at least one replicate showed undetermined Cq values (R^2^=0.99).

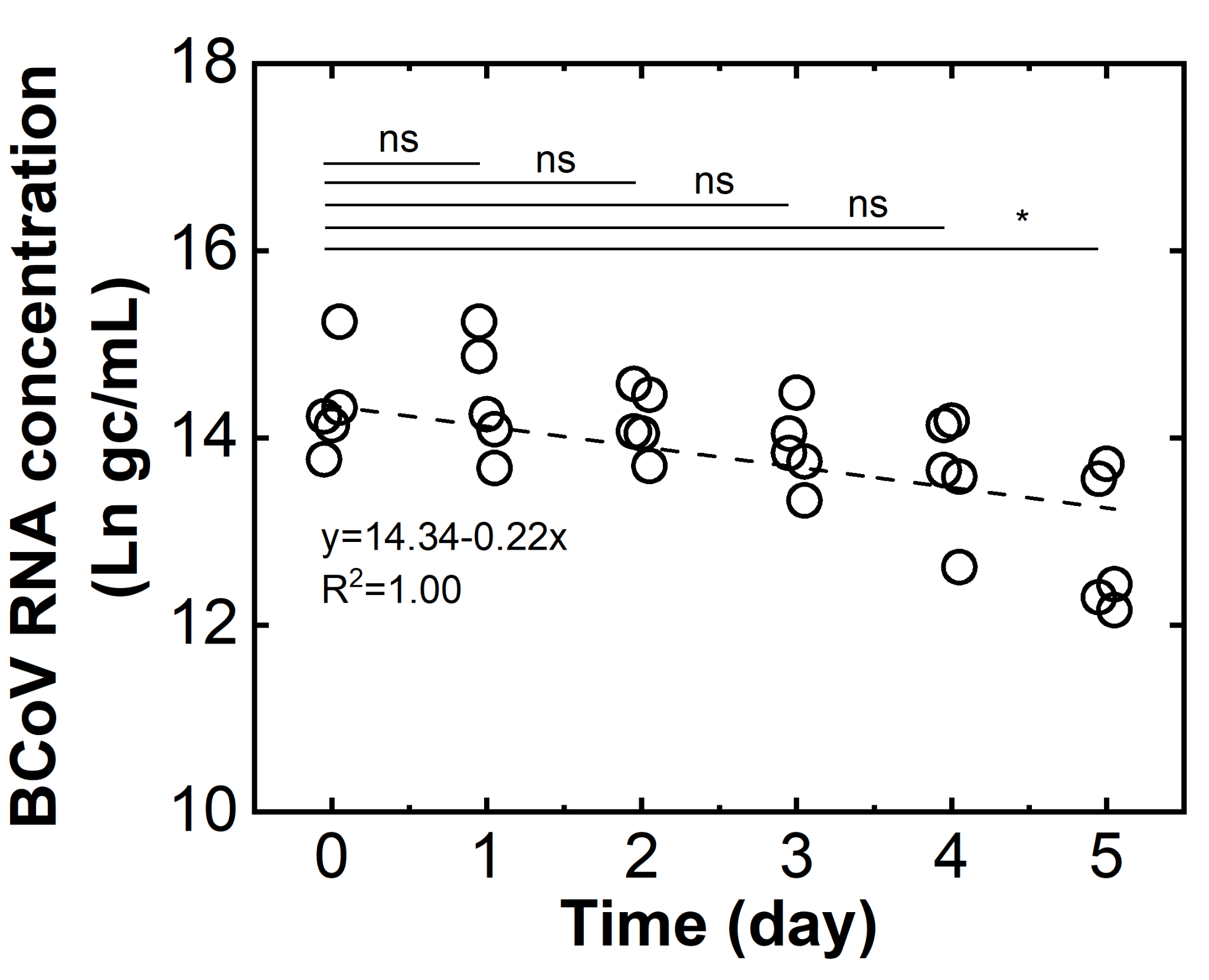

**Fig. S3.** BCoV RNA persistence in sewage samples at 25℃. The regression line was determined using the first-order decay kinetics. Statistical analyses of two groups of BCoV RNA concentrations were conducted by Mann-Whitney U Test (*: p<0.05 and ns: no significant difference).

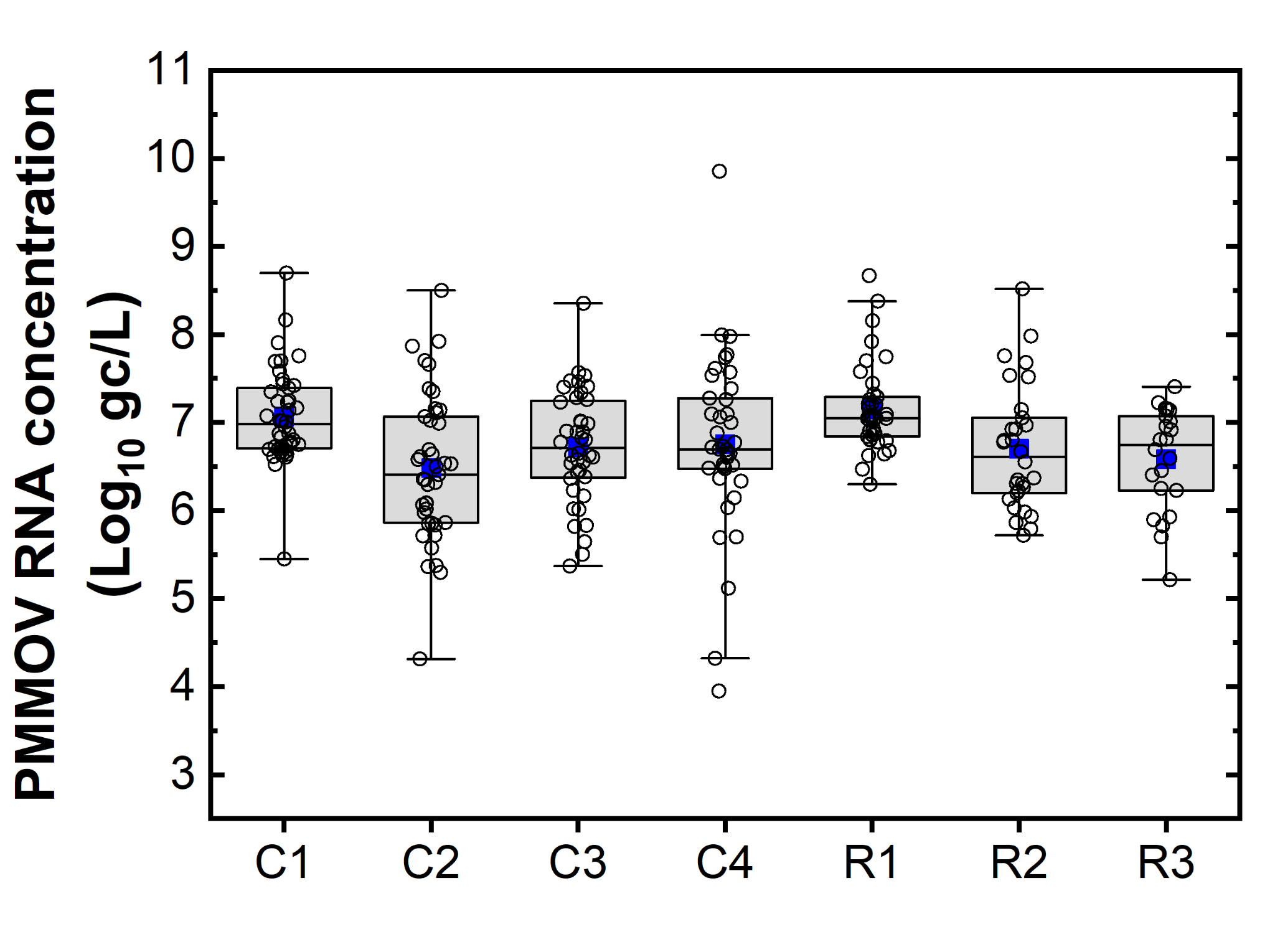

**Fig. S4.** PMMOV RNA concentrations in sewage samples collected from seven sewersheds. Whiskers of box plot indicate a 3×interquartile range. Blue rectangular solid boxes represent medians.

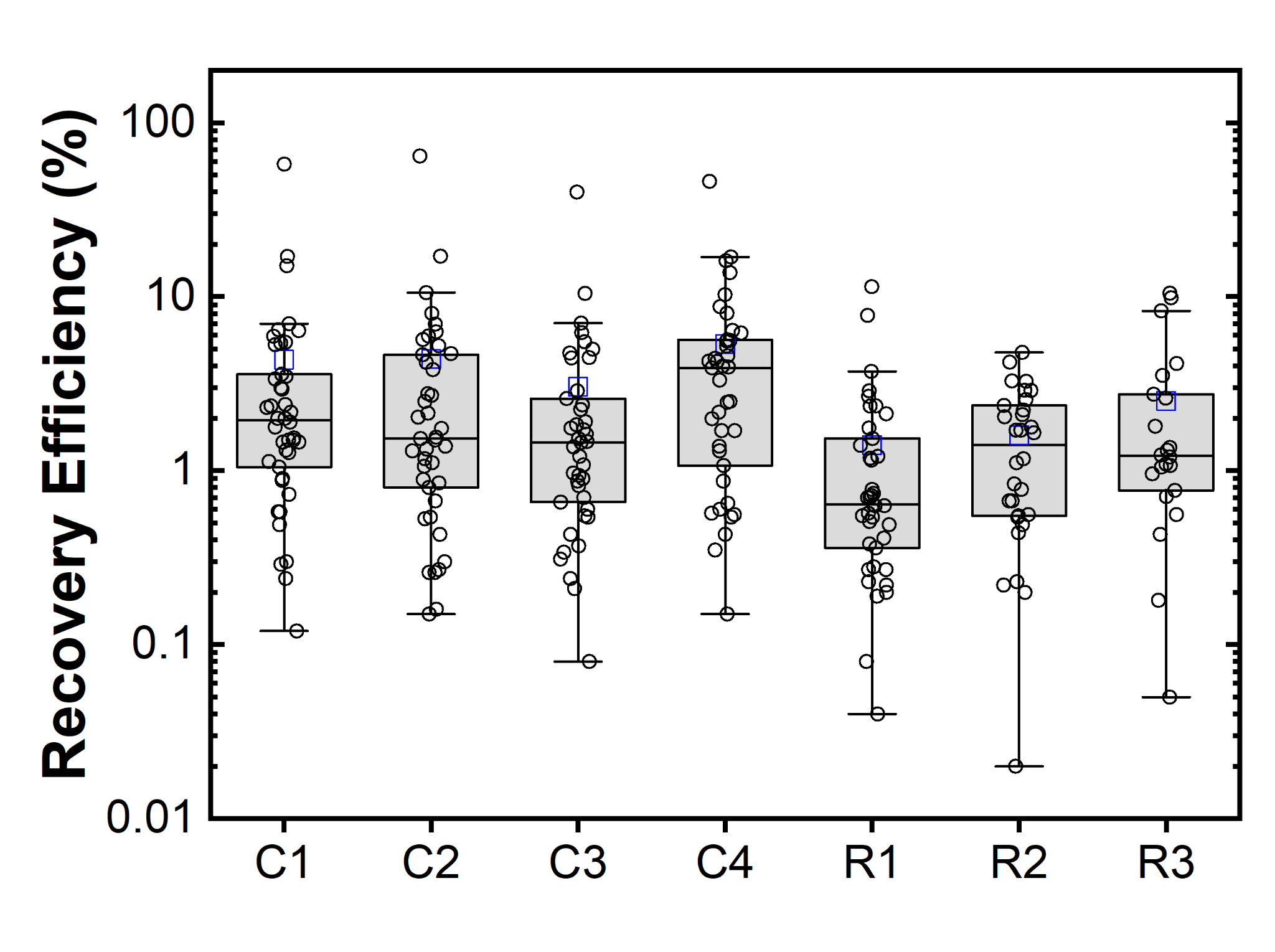

**Fig. S5.** Recovery efficiency of BCoV from sewage samples collected from seven sewersheds. Whiskers of box plot indicate a 3×interquartile range. Blue rectangular solid boxes represent medians.

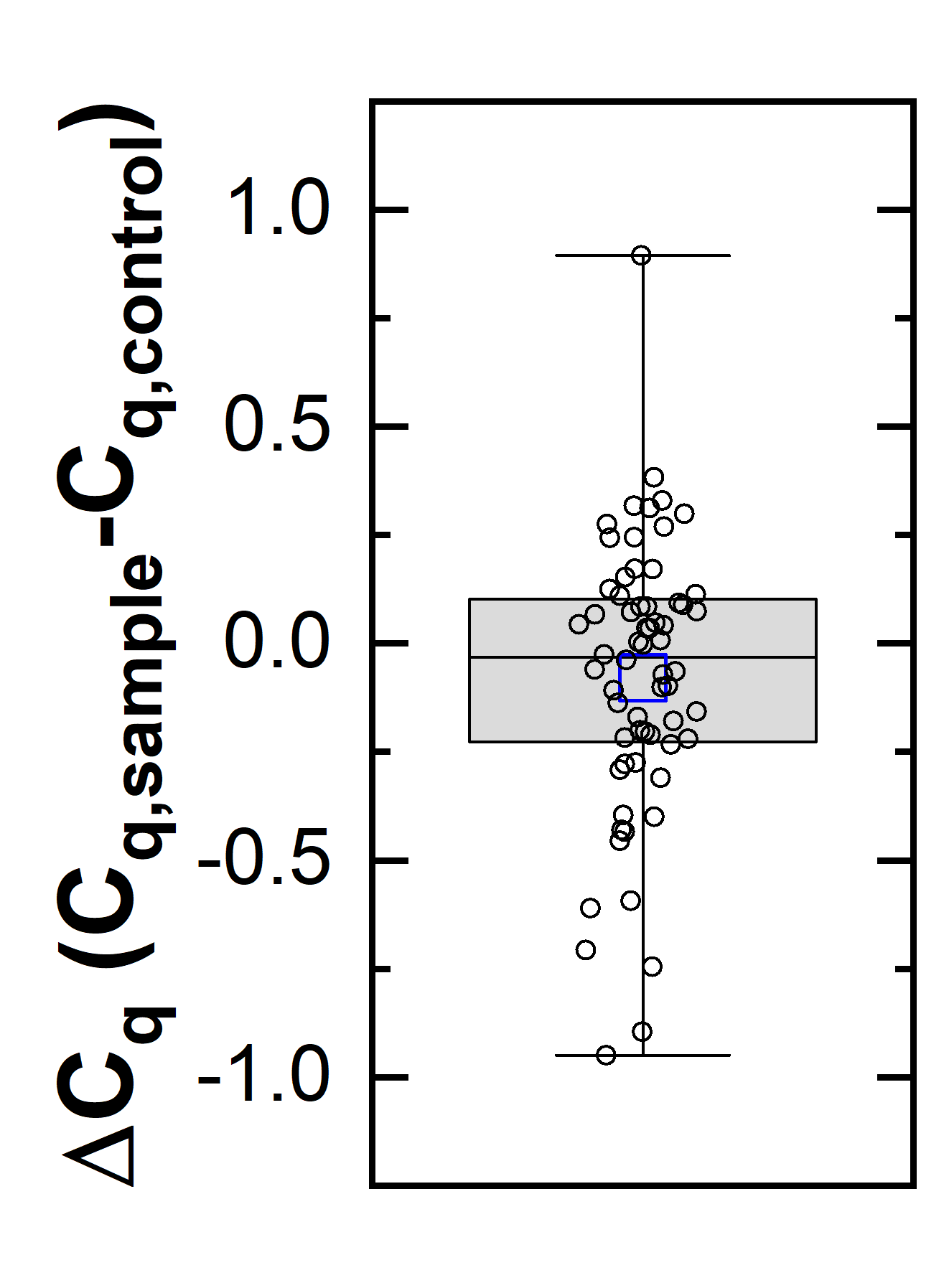

**Fig. S6.** Inhibition test with TV RNA. Whiskers of boxplot indicate a 1.5×interquartile range. Blue rectangular solid boxes represent medians.

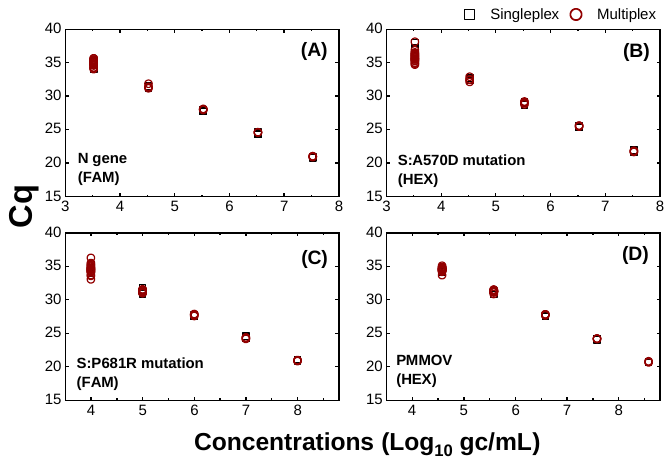

**Fig. S7.** Verification of multiplex RT-qPCR by comparing Cq values with corresponding singleplex RT-qPCR. The first set of multiplex RT-qPCR was used for (A) the N gene (FAM) and (B) S:A570D mutation (HEX) detection. The second set of multiplex RT-qPCR was applied for (C) the S:P681R mutation (FAM) and (D) PMMOV (HEX) detection.

**Table S7.** Summary of Spearman’s rank correlation coefficients determined from comparisons between N/PMMOV and three epidemiological factors: daily COVID-19 cases, 7-day average COVID-19 cases, and relative infection COVID-19 cases

| Site | Factors^2)^ | Time lag between clinical epidemiology and WBE data^1)^ | | | | | | | | | | | | | | |
| --- | --- | --- | --- | --- | --- | --- | --- | --- | --- | --- | --- | --- | --- | --- | --- | --- |
|  |  | -7 | -6 | -5 | -4 | -3 | -2 | -1 | 0 | 1 | 2 | 3 | 4 | 5 | 6 | 7 |
| C1 | D | 0.27 | 0.31 | 0.37 | 0.38 | 0.33 | 0.32 | 0.32 | 0.28 | 0.36 | 0.38 | 0.37 | 0.43 | 0.42 | 0.36 | 0.34 |
|  | A | 0.58 | 0.61 | 0.64 | 0.65 | 0.64 | 0.63 | 0.60 | 0.58 | 0.59 | 0.62 | 0.65 | 0.67 | 0.67 | 0.65 | 0.59 |
|  | R | 0.61 | 0.63 | 0.65 | 0.67 | 0.67 | 0.66 | 0.65 | 0.64 | 0.64 | 0.67 | 0.70 | 0.71 | 0.72 | 0.70 | 0.65 |
| C2 | D | 0.13 | 0.19 | 0.20 | 0.21 | 0.19 | 0.16 | 0.17 | 0.22 | 0.17 | 0.18 | 0.19 | 0.22 | 0.24 | 0.20 | 0.14 |
|  | A | 0.36 | 0.35 | 0.33 | 0.34 | 0.38 | 0.41 | 0.41 | 0.38 | 0.36 | 0.34 | 0.35 | 0.35 | 0.32 | 0.30 | 0.28 |
|  | R | 0.35 | 0.35 | 0.35 | 0.36 | 0.39 | 0.42 | 0.43 | 0.44 | 0.43 | 0.41 | 0.41 | 0.40 | 0.37 | 0.34 | 0.32 |
| C3 | D | -0.02 | 0.07 | 0.00 | -0.07 | -0.03 | -0.11 | -0.08 | 0.02 | 0.03 | 0.07 | 0.09 | 0.10 | 0.07 | 0.04 | 0.08 |
|  | A | 0.05 | 0.02 | -0.02 | -0.05 | -0.03 | -0.02 | 0.01 | 0.06 | 0.09 | 0.14 | 0.16 | 0.14 | 0.14 | 0.15 | 0.16 |
|  | R | 0.06 | 0.04 | 0.01 | -0.02 | -0.02 | 0.01 | 0.03 | 0.07 | 0.11 | 0.14 | 0.15 | 0.14 | 0.14 | 0.15 | 0.17 |
| C4 | D | 0.11 | 0.09 | 0.09 | 0.10 | 0.13 | 0.16 | 0.20 | 0.16 | 0.11 | 0.16 | 0.17 | 0.23 | 0.24 | 0.15 | 0.09 |
|  | A | 0.20 | 0.21 | 0.26 | 0.30 | 0.31 | 0.32 | 0.32 | 0.30 | 0.31 | 0.32 | 0.33 | 0.35 | 0.36 | 0.36 | 0.33 |
|  | R | 0.25 | 0.25 | 0.27 | 0.30 | 0.32 | 0.32 | 0.32 | 0.32 | 0.31 | 0.33 | 0.36 | 0.39 | 0.40 | 0.40 | 0.38 |
| R1 | D | 0.23 | 0.14 | 0.08 | 0.17 | 0.17 | 0.26 | 0.32 | 0.31 | 0.36 | 0.31 | 0.29 | 0.27 | 0.28 | 0.28 | 0.27 |
|  | A | 0.31 | 0.31 | 0.32 | 0.33 | 0.35 | 0.41 | 0.47 | 0.50 | 0.52 | 0.52 | 0.52 | 0.53 | 0.50 | 0.48 | 0.46 |
|  | R | 0.32 | 0.32 | 0.32 | 0.35 | 0.38 | 0.43 | 0.47 | 0.51 | 0.52 | 0.52 | 0.53 | 0.53 | 0.53 | 0.53 | 0.52 |
| R2 | D | 0.15 | 0.21 | 0.21 | 0.07 | 0.16 | 0.13 | 0.19 | 0.23 | 0.15 | 0.15 | 0.14 | 0.15 | 0.18 | 0.18 | 0.17 |
|  | A | 0.20 | 0.27 | 0.31 | 0.32 | 0.34 | 0.31 | 0.31 | 0.33 | 0.33 | 0.36 | 0.34 | 0.32 | 0.32 | 0.31 | 0.29 |
|  | R | 0.17 | 0.22 | 0.27 | 0.29 | 0.32 | 0.32 | 0.33 | 0.36 | 0.37 | 0.37 | 0.37 | 0.36 | 0.36 | 0.36 | 0.34 |
| R3 | D | -0.05 | 0.11 | 0.08 | 0.05 | -0.03 | -0.11 | -0.10 | -0.09 | 0.04 | 0.12 | 0.12 | -0.09 | -0.11 | -0.20 | -0.13 |
|  | A | 0.15 | 0.14 | 0.08 | 0.03 | 0.03 | 0.05 | 0.07 | 0.05 | 0.00 | -0.02 | -0.07 | -0.03 | -0.02 | 0.02 | 0.03 |
|  | R | 0.06 | 0.08 | 0.08 | 0.05 | 0.04 | 0.06 | 0.08 | 0.07 | 0.03 | 0.00 | -0.03 | -0.03 | -0.02 | 0.02 | 0.03 |

1) A negative time lag means WBE signals come earlier than clinical epidemiology.
2) D, A, and R indicate daily COVID-19 cases, 7-day average COVID-19 cases, and relative infection COVID-19 cases, respectively.

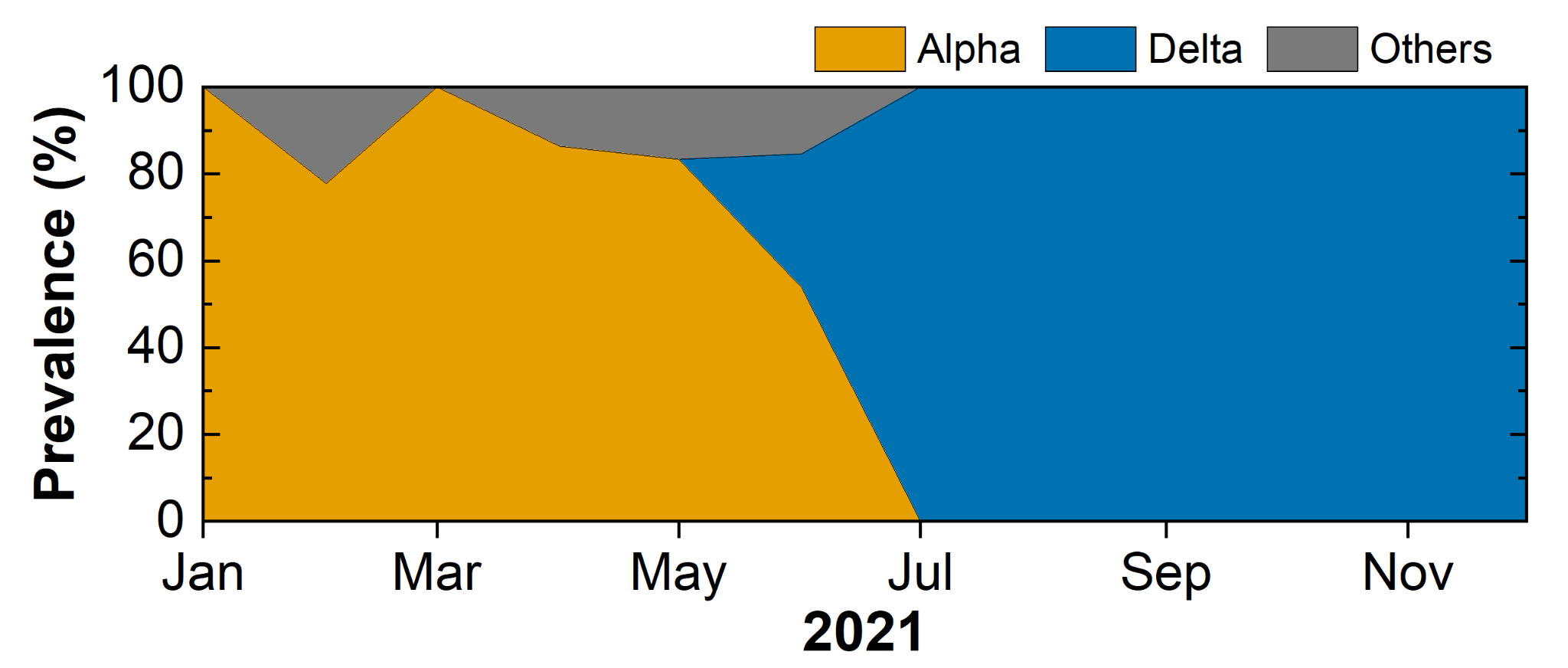

**Fig. S8.** SARS-CoV-2 variant prevalence in Champaign County. The variant dynamics was determined based on genome sequences deposited to GISAD (n=138).

**Table S8**. Demographic information of seven sewersheds

| Site | Area (km^2^) | Population | Rate | | | | Hispanic | Under age 18 | Housing vacancy |
| --- | --- | --- | --- | --- | --- | --- | --- | --- | --- |
|  |  |  | White | Black | Asian | Two or more/Other |  |  |  |
| C1 | 0.09 | 1675 | 39.4% | 6.1% | 44.8% | 9.7% | 9.4% | 3.8% | 9.8% |
| C2 | 0.51 | 1260 | 33.5% | 45.9% | 2.7% | 17.9% | 9.8% | 31.5% | 10.5% |
| C3 | 0.37 | 853 | 30.9% | 37.4% | 7.7% | 24.0% | 16.2% | 26.9% | 5.7% |
| C4 | 0.20 | 859 | 32.2% | 55.9% | 2.2% | 9.7% | 3.9% | 31.9% | 6.4% |
| R1 | 1.09 | 2402 | 59.0% | 20.9% | 0.7% | 19.4% | 16.3% | 24.8% | 10.1% |
| R2 | 1.70 | 2160 | 50.5% | 13.8% | 1.6% | 34.2% | 31.6% | 28.0% | 12.6% |
| R3 | 0.73 | 1088 | 41.1% | 41.7% | 0.9% | 16.3% | 7.9% | 36.0% | 11.1% |
| All seven sewersheds combined | 4.69 | 10297 | 44.2% | 23.6% | 12.9% | 19.3% | 15.5% | 22.4% | 10.1% |
| Champaign County | 2582.20 | 205865 | 62.7% | 13.9% | 11.9% | 11.5% | 8.1% | 19.2% | 9.3% |
